# Protein associations and protein–metabolite interactions with depressive symptoms and the p-factor

**DOI:** 10.1101/2024.10.30.24316418

**Authors:** Alyce M Whipp, Gabin Drouard, Richard J Rose, Lea Pulkkinen, Jaakko Kaprio

## Abstract

Despite increasing mental health problems among young people, few studies have examined associations between plasma proteins and mental health, and interactions between proteins and metabolites in association with mental health problems remain underexplored. In 730 twins, we quantified associations between plasma proteins measured at age 22 with 21 indicators representing either depressive symptoms or the p-factor, collected from questionnaires and interviews completed by different raters (e.g., self-report, teachers) through adolescence to young adulthood (12 to 22 years), and tested for interactions with metabolites. We found 47 proteins associated with depressive symptoms or the p-factor (FDR<0.2), 9 being associated with both. Two proteins, contactin-1 and mast/stem cell growth factor receptor kit, positively interacted with valine levels in explaining p-factor variability. In conclusion, our study demonstrates strong associations between plasma proteins and mental health and provides evidence for proteome– metabolome interactions in explaining higher levels of mental health problems.

## Introduction

With mental health problems on the rise^1^, and the poor success rates of existing treatments for mental health disorders like depression^2^, we continue to need to improve our understanding of the biological mechanisms involved in these disorders. The omics revolution has brought the ability to examine mechanisms of a health problem from the genomic and epigenomic levels, and more recently from the gut microbiome, metabolomic and proteomic levels. As these omics technologies are now being used in more and more investigations of mental health^3–5^, we are starting to have more pieces of the puzzle, however, these disorders are heterogeneous and complex and we continue to seek more clarity, including by using multi-omic approaches.

The proteome and metabolome refer to the total set of proteins and metabolites, respectively, within a biological system, most commonly measured in blood. While the proteome and metabolome may differ in nature, they are both influenced by genetic and environmental factors^6,7^ and are dynamic, making them optimal resources for biomarker investigations. Combining proteomic data with metabolomic data would allow for refining our understanding of the link between the beginning of the biologic pathway (the genetics) and the end (the metabolomics) for a health problem, since the proteome is an intermediary. Additionally, the different levels of investigation that multi-omics allow for could also provide more clues about the lesser-known triggers to poor mental health such as inflammatory disorders, infections, or metabolic conditions^8,9^. For example, in a recent multi-omic investigation on body mass index (BMI) by our group, combining proteomics, metabolomics, transcriptomics, and polygenic risk scores allowed us to show that the associations between plasma proteins and changes in BMI during adolescence were characterized by common metabolic etiologies^10^. Regarding mental health, to our knowledge, multi-omic approaches are scarce, and we are unaware of previous studies that have looked at interactions between the proteome and metabolome. To date, even single-omic approaches to mental health have been limited in the literature and have involved a single outcome, as has been demonstrated in two recent studies using the Strengths and Difficulties Questionnaire (SDQ)^11^ and the p-factor^12^. Furthermore, while there have been a few large-scale studies that have investigated mental health with proteomics data, using for example UK Biobank data^13^, these studies were performed on older adults only. Whether the association of the plasma proteome with depressive symptoms and the p-factor differs by age, or by rater, has therefore not been investigated to the best of our knowledge.

Previously, our group has investigated the metabolomics of mental health, including depressive symptoms and aggressive behavior, in young adult Finnish twins. We found two branched-chain amino acids (BCAAs) -- valine and leucine -- that were (negatively) associated with depressive symptoms in our FinnTwin12 cohort^14^. The trend in these associations were consistent in meta-analysis across multiple ages, raters, and instruments of depressive symptoms. Additionally, we found a ketone body (3-hydroxybutyrate) that was (negatively) associated with aggressive behavior, and was suggestively replicated in a Dutch twin sample^15^. Furthermore, biomarkers for the p-factor were also sought, but no additional metabolites from the above findings were identified^14^. The p-factor, or psychopathology factor, initially characterized by Caspi et al.^16^, aggregates internalizing disorders (such as depression), externalizing disorders (such as aggression/conduct problems), and thought disorders (such as schizophrenia) into a higher-order combined dimension, which has shown a common underlying genetics^17^, despite the heterogeneity in the disorders. Our group has recently acquired proteomic data on the same cohort as the metabolomic analyses were performed, and thus have the unique opportunity to use a multi-omic approach to further tease out our previous findings.

Thus, we first aim to investigate, in an exploratory way, proteomic associations with depressive symptoms and the p-factor in our FinnTwin12 cohort (Figure 1). We used a total of 21 outcomes that describe depressive symptoms (including measures of major depressive disorder (MDD) and summary scores of general depressive symptoms) and comprise the p-factor (combining internalizing and externalizing problems) from different raters and that were reported at different ages, from childhood to young adulthood. Furthermore, for significant protein associations, we aim to investigate possible protein–metabolite interactions with the significant BCAA and 3-hydroxybutyrate metabolite associations found in the previous FinnTwin12 investigations. This study plan was pre-registered at Open Science Framework (OSF) in May 2023 (<u>osf.io/kc9hw</u>).

**Figure 1:**
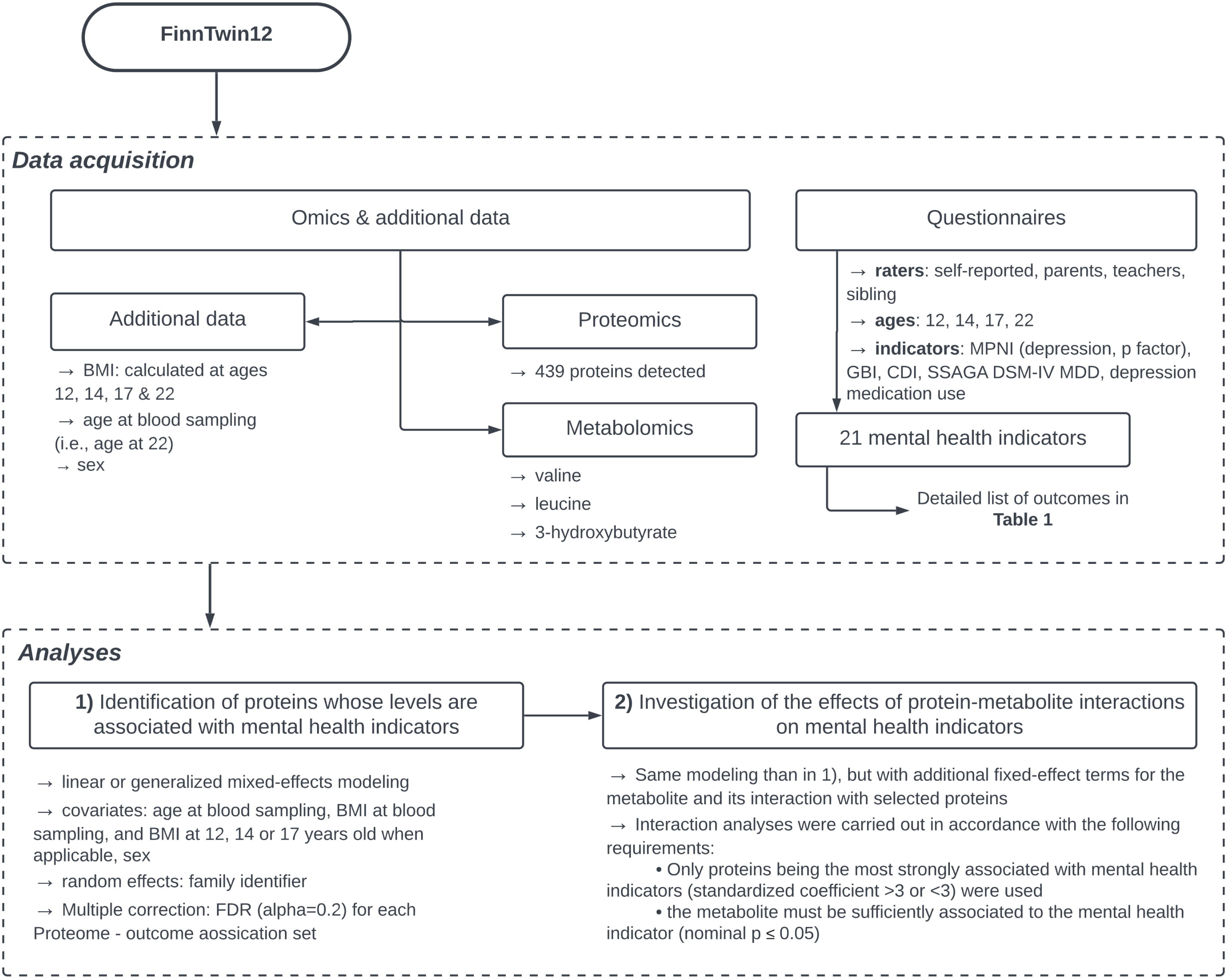
Overall study workflow.

## Results

We first modeled the 21 different mental health indicators (Table 1) as dependent variables and age at blood sampling, sex, and BMI as covariates. A description of the mental health indicators and their intricate relationships is presented in Table 1 and Figure 2, respectively. We found 68 significant associations between proteins and mental health indicators at an FDR ≤0.2 level (Table 2; 628 associations with nominal p<0.05, see Supplemental Table 1). Altogether, significant associations involved 47 unique proteins, of which 17 were associated with two or more mental health indicators. Of the 68 associations, 24 were with depressive symptom variables (CDI-14 (n=1; effect size −0.65), GBI-22 (n=4; effect sizes ranging - 0.57 to 0.61), and MPNI-s14d (n=19; effect sizes ranging −0.05 to 0.05)) and 44 with p-factor variables (MPNI-pz (n=11; effect sizes ranging −0.18 to 0.13), MPNI-s17p (n=11; effect sizes ranging −1.34 to 1.33), and MPNI-s14p (n=22; effect sizes ranging −0.99 to 1.08)). Some proteins were significantly associated with both depressive symptoms and p-factor variables (9 proteins), while some were significant only with depressive symptoms (13 proteins) or the p-factor (25 proteins)(Figure 3). Additionally, 58 of the associations were negative, 10 positive. The strongest associations were found with the p-factor, with the highest statistical significance between: IgGFc-binding protein with MPNI-pz (t-value=-4.81; nominal p=1.9e-6) and MPNI-co14d (t-value=-4.0; nominal p=5.9e-5), complement C4-b with MPNI-s14d (t-value=4.1; nominal p=4.4e-5), Fibulin-1 with MPNI-s17d (t-value=-4.1; nominal p=5.0e-5), and Monocyte Differentiation Antigen CD14 with MPNI-pz (t-value=4.0; nominal p=7.5e-5)(Table 2).

**Table 1:** Description of 21 mental health indicators in participants with proteomic data **Legend:** The 21 mental health indicators were calculated from questionnaires completed by different raters at different ages during adolescence and young adulthood. The number of participants with proteomic data was 730. The symbol * denotes categorical indicators; others are continuous. N: Number of participants with proteomic data who also had non-missing information for each mental health indicator. sd: standard deviation.

| Variable name | Instrument/Scale | Age at report | Rater | N | Mean | SD | Range | Skewness | Kurtosis |
| --- | --- | --- | --- | --- | --- | --- | --- | --- | --- |
| MPNI-p12d | MPNI - depression | 12 | parent(s) | 694 | 0.75 | 0.4 | 0 - 2.4 | 0.8 | 0.5 |
| MPNI-t12d | MPNI - depression | 12 | teacher | 704 | 0.64 | 0.5 | 0 - 2.8 | 1.0 | 1.2 |
| MPNI-t14d | MPNI - depression | 14 | teacher | 565 | 0.53 | 0.5 | 0 - 2.4 | 1.0 | 1.0 |
| MPNI-s14d | MPNI - depression | 14 | self-report | 692 | 0.66 | 0.4 | 0 - 2.4 | 0.6 | 0.3 |
| MPNI-co14d | MPNI - depression | 14 | co-twin | 695 | 0.61 | 0.4 | 0 - 2.2 | 0.6 | 0.2 |
| MPNI-s17d | MPNI - depression | 17 | self-report | 655 | 0.73 | 0.7 | 0 - 3.0 | 0.8 | 0.1 |
| MPNI-co17d | MPNI - depression | 17 | co-twin | 650 | 0.67 | 0.6 | 0 - 3.0 | 0.7 | 0.0 |
| CDI-14 | CDI | 14 | self-report | 704 | 6.25 | 5.1 | 0 - 40 | 1.7 | 5.2 |
|  | SSAGA DSM-IV MDD |  |  |  |  |  |  |  |  |
|  | lifetime depressive |  |  |  |  |  |  |  |  |
| MDD-14 | symptoms | 14 | self (interview) | 726 | 0.23 | 0.61 | 0 - 5.0 | 3.63 | 18.1 |
|  | SSAGA DSM-IV MDD |  |  |  |  |  |  |  |  |
| MDD-22 | diagnosis, Y/N | 22 | self (interview) | 730 | cases: 14% |  |  |  |  |
| GBI-17 | GBI | 17 | self-report | 663 | 4.90 | 4.8 | 0 - 30 | 1.6 | 3.3 |
| GBI-22 | GBI | 22 | self-report | 723 | 4.49 | 4.6 | 0 - 29 | 1.8 | 4.5 |
|  | Using depression |  |  |  |  |  |  |  |  |
| medication-22d | medication, Y/N | 22 | self (interview) | 730 | cases: 3% |  |  |  |  |
| MPNI-p12p | MPNI - pfactor | 12 | parent(s) | 688 | 16.22 | 7.9 | 0 - 49 | 0.8 | 0.8 |
| MPNI-t12p | MPNI - pfactor | 12 | teacher | 688 | 15.47 | 11.4 | 0 - 61 | 1.0 | 0.9 |
| MPNI-t14p | MPNI - pfactor | 14 | teacher | 554 | 11.17 | 9.3 | 0 - 50 | 1.4 | 1.7 |
| MPNI-s14p | MPNI - pfactor | 14 | self-report | 682 | 16.38 | 6.8 | 2 - 44 | 0.6 | 0.5 |
| MPNI-co14p | MPNI - pfactor | 14 | co-twin | 675 | 16.53 | 8.2 | 1 - 46 | 0.6 | 0.3 |
| MPNI-s17p | MPNI - pfactor | 17 | self-report | 631 | 19.22 | 6.5 | 3 - 41 | 0.4 | 0.0 |
| MPNI-co17p | MPNI - pfactor | 17 | co-twin | 627 | 19.13 | 7.8 | 3 - 46 | 0.4 | -0.3 |
|  | MPNI - pfactor (z-scored |  |  |  |  |  |  |  |  |
|  | score of all 7 individual |  |  |  |  |  |  |  |  |
| MPNI-pz | pfactor variables) | - | - | 721 | -0.03 | 1.0 | -2.2 - 4.3 | 0.6 | 0.4 |

**Figure 2:**
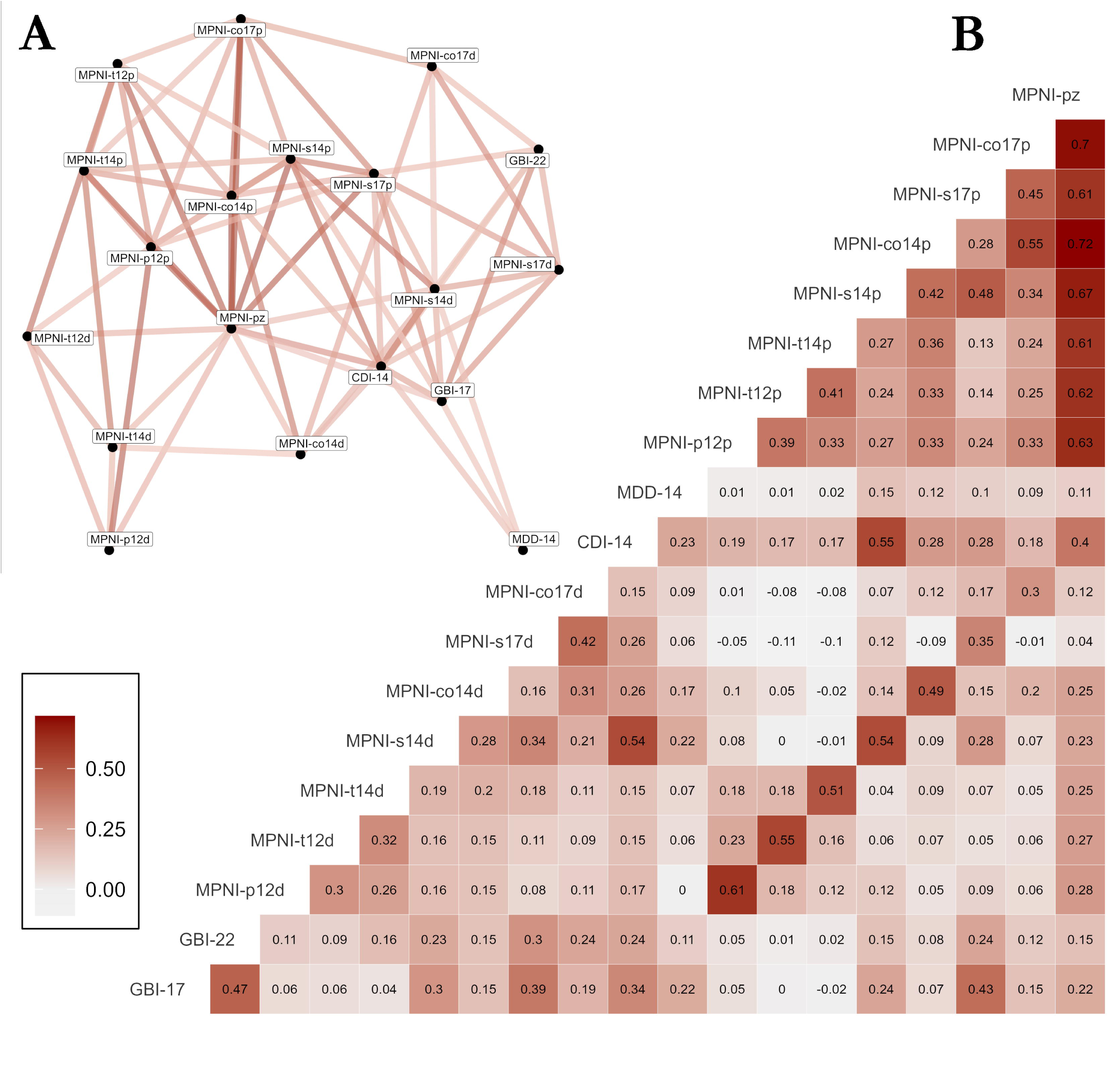
Pairwise Pearson correlations between continuous mental health indicators **Legend:** Pairwise Pearson correlations are presented in two different formats: **(A)** a graph connecting mental health indicators if the pairwise Pearson correlation exceeds 0.2 in absolute value, and **(B)** a correlation matrix.

**Table 2:**
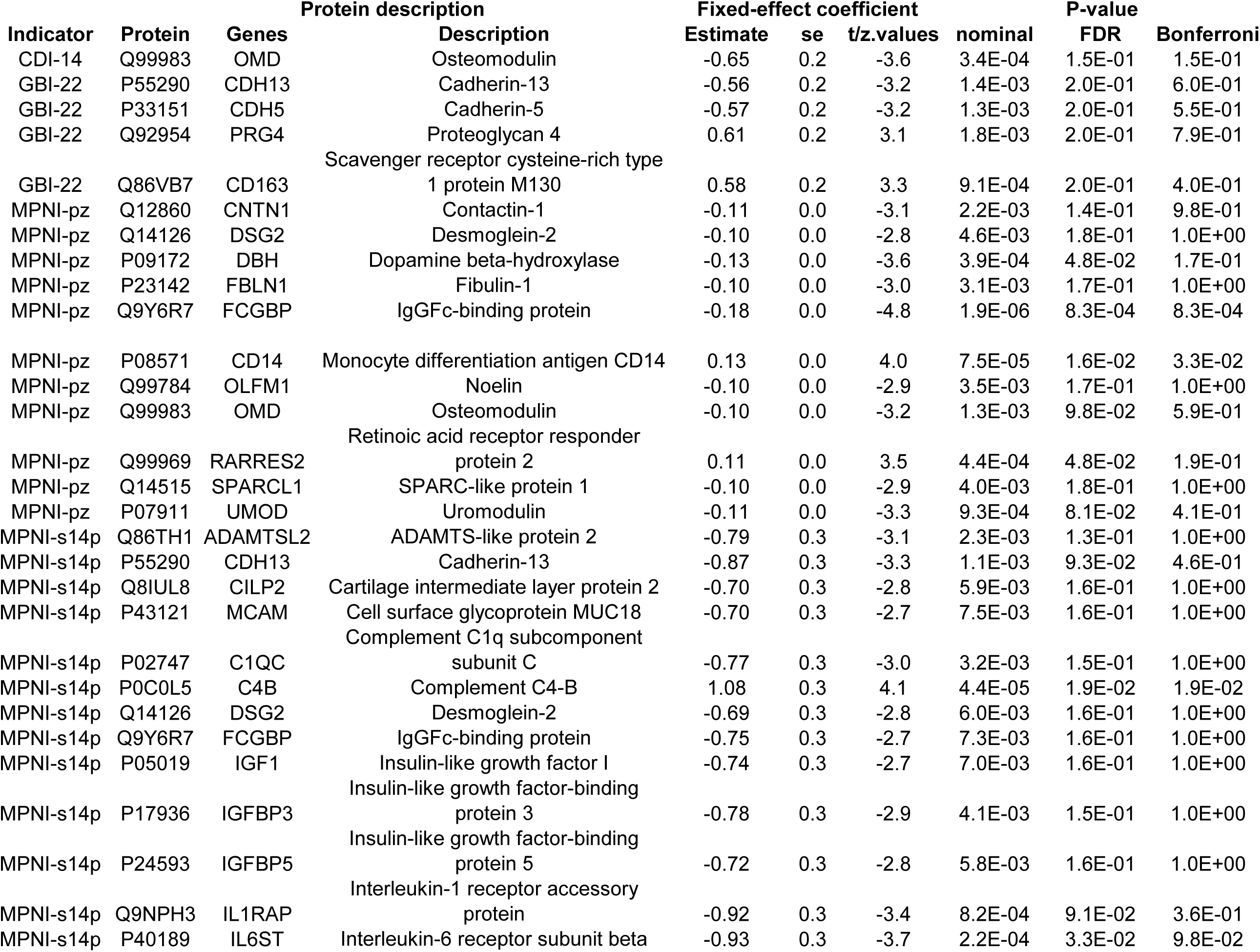
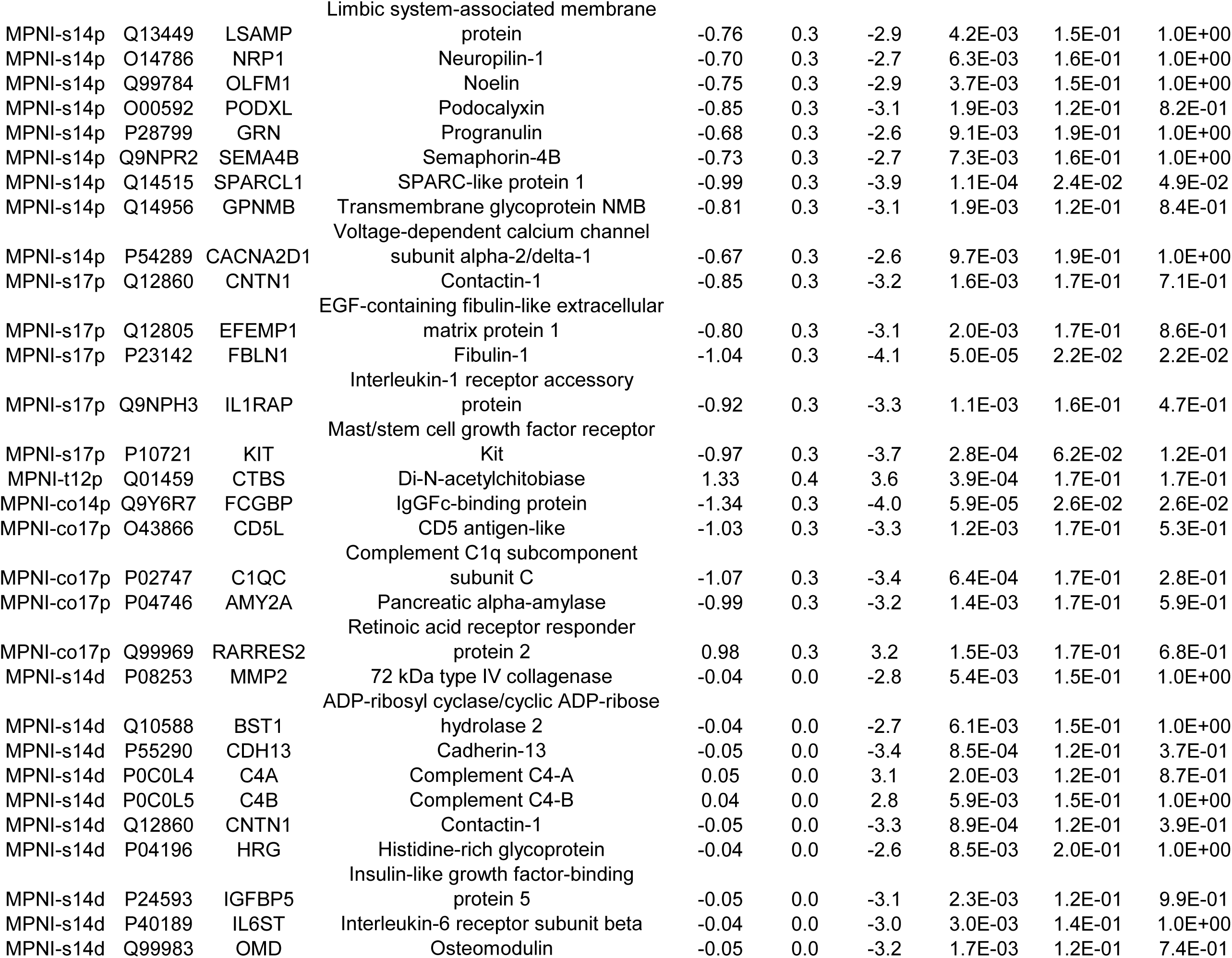
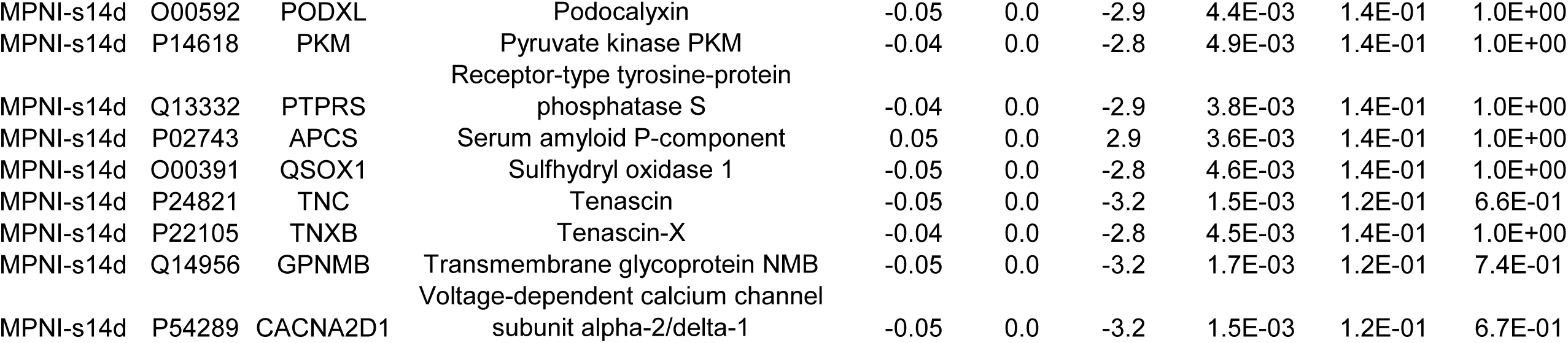
Associations between protein abundance and mental health indicators. **Legend:** Multiple correction was performed at the level of each mental health indicator using Bonferroni or FDR correction. Only associations for which FDR-adjusted p-value is below 0.2 are presented. FDR: False Discovery Rate. se: standard error.

**Figure 3:**
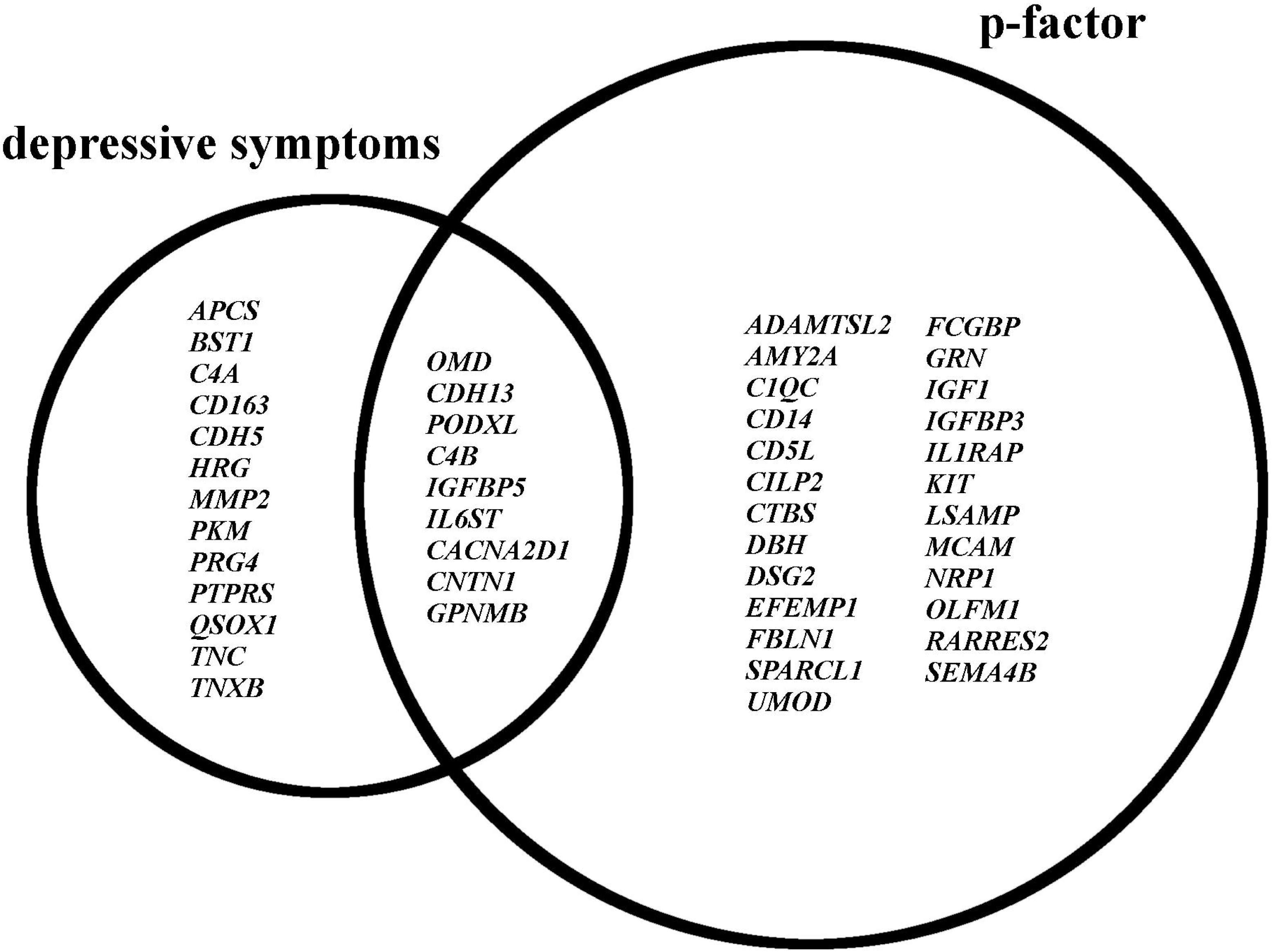
Venn diagram of protein-coding genes **Legend:** The genes encoding the identified proteins were partitioned according to whether their encoded proteins were associated only with depressive symptoms (CDI, MPNId, GBI), only with the p-factor (across raters and age groups), or with both. See Table 2 for protein descriptions.

In follow-up analyses, of those 47 proteins with significant associations with mental health indicators, those that were strongly associated with mental health (see Figure 1) were investigated for protein–metabolite associations with mental health (the metabolite was also assessed to be highly significantly associated with mental health; see Figure 1). Thus, 7 interactions were tested. Of these, 2 protein–metabolite interactions were found with MPNI-s17p: Mast/stem cell growth factor receptor Kit and Valine (interaction term p=0.01), and Contactin-1 and Valine (interaction term p=0.04)(Table 3). To further illustrate the protein–metabolite interactions with mental health, the z-scored levels of these two valine-interacting proteins were then plotted with the self-reported p-factor values at age 17 (Figure 4). Stratification by the first and last quartile of valine was performed to visualize shifts in protein associations with mental health. In participants with high levels of valine, the deleterious effects of the Contactin-1 or Mast/stem cell growth factor receptor kit on mental health are lower independent of valine’s effect alone.

**Table 3:** Interactions between selected protein abundance and metabolites. **Legend:** Interactions between proteins and metabolites were tested for proteins identified in Table 2 with standardized coefficients above 3 in absolute value, and if the metabolite was previously associated with the mental health indicator. FDR: False Discovery Rate. se: standard error.

| outcome | Protein |  | Metabolite | Protein estimate |  |  |  | Metabolite estimate |  |  |  | Protein:Metabolite interaction |  |  |  |
| --- | --- | --- | --- | --- | --- | --- | --- | --- | --- | --- | --- | --- | --- | --- | --- |
|  | UniProt | Gene |  | coef | se | t-value | p | coef | se | t-value | p | coef | se | t-value | p |
| MPNI-s17p | P10721 | KIT | Valine | 35.1 | 13.1 | 2.7 | 0.01 | -2.6 | 1.1 | -2.4 | 0.02 | -2.3 | 0.8 | -2.7 | **0.01** |
| MPNI-s17p | P23142 | FBLN1 | Valine | 17.2 | 13.3 | 1.3 | 0.20 | -2.1 | 1.1 | -1.9 | 0.06 | -1.1 | 0.8 | -1.4 | 0.17 |
| MPNI-s17p | Q12805 | EFEMP1 | Valine | 21.7 | 12.6 | 1.7 | 0.09 | -2.4 | 1.1 | -2.2 | 0.03 | -1.4 | 0.8 | -1.8 | 0.08 |
| MPNI-s17p | Q12860 | CNTN1 | Valine | 28.4 | 14.4 | 2.0 | 0.05 | -2.2 | 1.1 | -2.0 | 0.04 | -1.8 | 0.9 | -2.0 | **0.04** |
| MPNI-s17p | Q9NPH3 | IL1RAP | Valine | 10.1 | 12.9 | 0.8 | 0.43 | -2.7 | 1.1 | -2.4 | 0.02 | -0.7 | 0.8 | -0.9 | 0.39 |
| MPNI-t12p | Q01459 | CTBS | Valine | 0.7 | 19.1 | 0.0 | 0.97 | -3.8 | 1.6 | -2.4 | 0.02 | 0.0 | 1.2 | 0.0 | 0.98 |
| MPNI-t12p | Q01459 | CTBS | 3-hydroxybutyrate | 0.5 | 6.0 | 0.1 | 0.94 | -0.9 | 0.5 | -2.0 | 0.05 | 0.1 | 0.4 | 0.1 | 0.89 |

**Figure 4:**
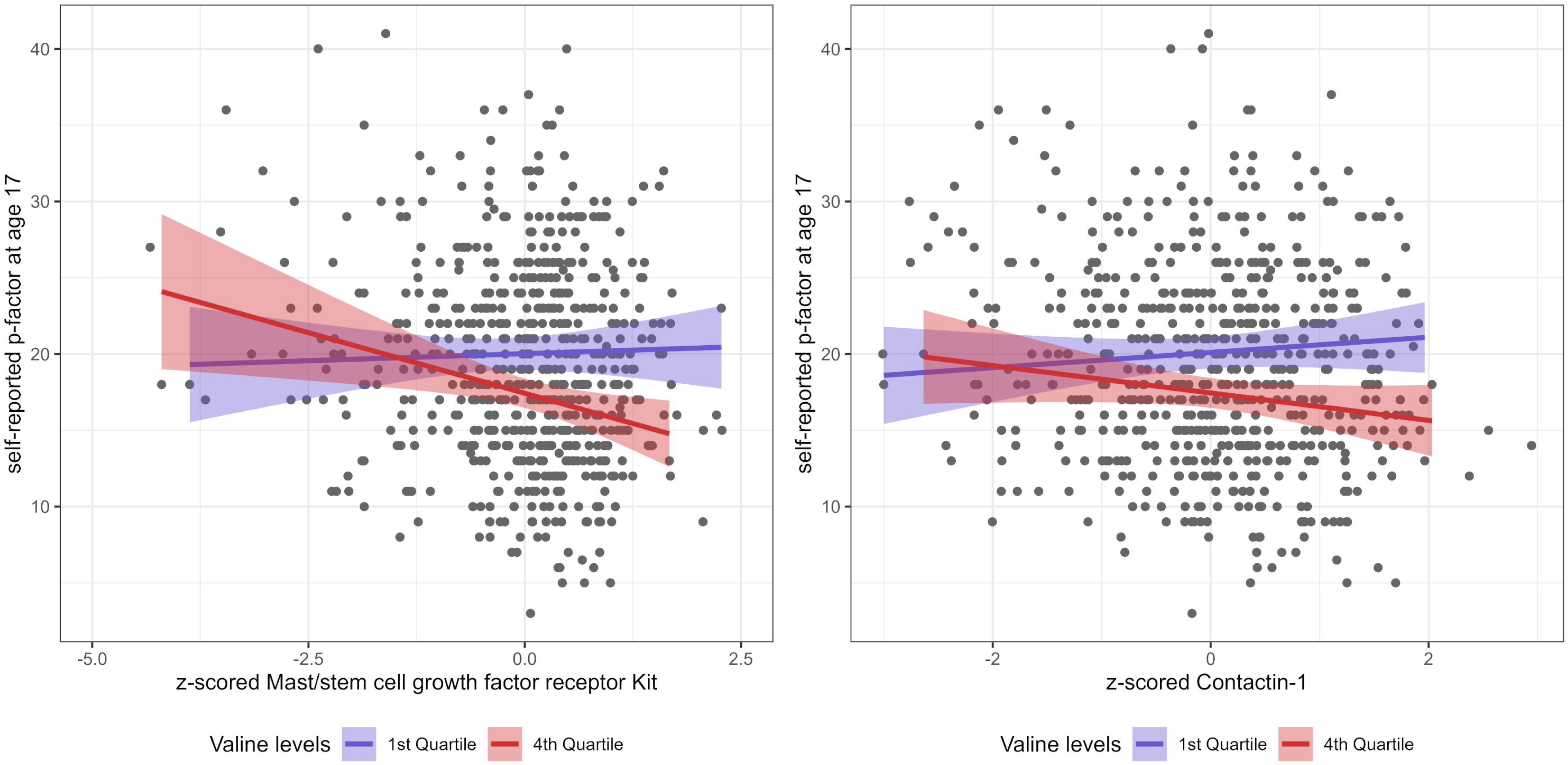
Graphical representation of significant protein–valine interactions **Legend:** The plot represents Mast/stem cell growth factor receptor Kit and Contactin-1 z-scored levels (x axis) in relation to the self-reported p-factor score at age 17 (y axis). Participants having non-missing valine data are plotted. The y∼x regression lines are shown for participants in the first or last quartile of valine levels.

## Discussion

In this exploratory multi-omic study of mental health, we found 47 proteins significantly associated with depressive symptoms or the p-factor in our FinnTwin12 cohort. Of these, 17 proteins were found to have more than one association across different ages, raters, and/or mental health indicators. The majority of protein associations were negative and involved the p-factor. Furthermore, we identified two protein–metabolite interactions associated with the p-factor; these involved valine (metabolite) and proteins Mast/stem cell growth factor receptor Kit and Contactin-1. These associations are not indications of diagnostic markers nor are they suggested to be causative, rather these proteins and associations suggest the best places to investigate new biological pathways of interest in better understanding mental health disorders.

Those proteins associated with mental health across different ages (e.g., Contactin-1, Cadherin-13), different raters (e.g., IgGFc-binding protein, Complement C1q subcomponent subunit C), and/or different mental health indicators (e.g., Podocalyxin, Complement C4-B) may lead to the most fruitful in-depth investigations of biological mechanisms, given the greater consistency of those associations. Some of the proteins (e.g., Contactin-1, IgGFc-binding protein) found in this more extended and refined analysis overlap with a previous investigation^12^, restricted to the p-factor only and using a different approach; this too supports the stability of these associations. Particularly noteworthy is that several of the identified proteins were associated with both the p-factor and depressive symptoms. As such, these proteins may reflect underlying shared information between the p-factor and depressive symptoms, which may indicate shared internalizing factors, since the p-factor aggregates internalizing factors along with externalizing factors (thought disorders are also included in the p-factor, though were not available in our dataset). Anxiety items and measures are not as abundant in our data collection, thus we did not include anxiety separately in this investigation although it could help clarify this overall p-factor– internalizing connection. However, phenotypic and genetic associations between externalizing factors and depression or depressive symptoms have been reported in the literature^18–20^. This suggests that the proteins associated with both the p-factor and depressive symptoms may not necessarily reflect only internalizing behaviors, but could potentially reflect a more complex picture that combines internalizing and externalizing factors.

Connecting the proteins with the most consistent associations with mental health shows that many have a high likelihood of yielding improved biological and treatment insights. For example, contactin-1 (associated with MPNI-pz, MPNI-s17p, MPNI-s14d, and has an interaction with valine) or its gene (CNTN1), involved in neuronal and oligodendrocyte development, has been shown to be associated with psychiatric illness (e.g., antidepressant treatment resistance, bipolar disorder) as well as neurodegenerative disease (e.g., Alzheimer’s and Parkinson’s)^21,22^. Additionally, CNTN1 has been associated in the national FinnGen database (www.fingen.fi) with 50,000+ cases of depression medication use and ‘psychiatric diseases’ among ethnic Finns^23^. In a recent large-scale proteomic analysis of older adults from the UK Biobank, the contactin-1 protein was also found to be negatively associated with depressive symptoms (log_10_(p)=10.6)^13^. This suggests that contactin-1 is a good biomarker for investigations of depression and related symptoms in all age groups.

Cadherin-13 (associated with GBI-22, MPNI-s14p, MPNI-s14d), a cell adhesion molecule, or its gene (CDH13) has also been shown in the literature to be associated with psychiatric illness (e.g., depression, substance abuse, attention-deficit/hyperactivity disorder) ^24–27^, as well as with psychiatric disorders among ethnic Finns in the FinnGen database^23^. Podocalyxin (associated with MPNI-s14p, MPNI-s14d), another protein involved in cell adhesion and transportation, or its gene (PODXL), has been shown to be involved with neural development, blood-brain barrier function, neuroinflammation, and neurodegenerative disease^28–30^, as well as with neurological and psychiatric disorders among ethnic Finns in the FinnGen database^23^. The gene (FCGBP) coding for IgGFc-binding protein (associated with MPNI-pz, MPNI-s14p, MPNI-co14p), a protein involved in immune response, has been shown to be associated with bipolar disorder and depression^31,32^, as well as with neurological and psychiatric disorders among ethnic Finns in the FinnGen database^23^. Another protein involved in immunity, complement C1q subcomponent subunit C (associated with MPNI-s14p, MPNI-co17p) or its gene (C1QC) has been associated with depression^33–35^, as well as with psychiatric disorders and depression among ethnic Finns in the FinnGen database^23^; furthermore, complement C4-B (associated with MPNI-s14p, MPNI-s14d) or its gene (C4B) is associated with white matter integrity, depression, and schizophrenia^36–38^, as well as with psychiatric disorders, depression, and schizotypal disorders among ethnic Finns in the FinnGen database^23^. Lastly, mast/stem cell growth factor receptor Kit (associated with MPNI-s17p and valine interaction), a protein involved in cell signaling, or its gene (KIT), have been associated with obsessive compulsive disorder, schizophrenia, and autism spectrum disorders^39,40^, as well as being associated in the FinnGen database with psychiatric disorders, depression, anxiety disorders, and schizotypal disorders^23^.

For mast/stem cell growth factor receptor Kit and contactin-1, a significant protein– metabolite interaction (metabolite: valine) was identified with the p-factor. While higher levels of contactin-1 or mast/stem cell growth factor receptor Kit were associated with higher p-factor scores at age 17, the effect sizes were reduced in participants with high levels of valine. To the best of our knowledge, these interactions have not previously been shown. Valine had previously been shown to be negatively associated with depressive symptoms, and approached significance with the p-factor^14^, and contactin-1 has been shown to be associated with depression phenotypes in mice^41,42^ and older adults from the UK Biobank^13^, as well as the previously mentioned psychiatric and neurodegenerative associations. Additionally, a study of essential amino acid (EAA) supplementation and protein expression in low physical activity older adults showed reduced expression of mast/stem cell growth factor receptor kit in relation to EAA supplementation^43^. Currently, however, the direct meaning of our found interactions cannot be clearly interpreted since how the two omic levels influence each other is poorly known. Additionally, our approach for testing for interactions was rather conservative in that we required independent association with both the metabolite (p<0.05) and the protein (|standardized coefficient| >3) before interaction testing, thus only 7 interactions were tested.

An additional point to raise with these findings involves the use of multiple raters and the frequent significant associations of proteins with self-reports of mental health. Self-reports from ages 14, 17, and 22 were among many of the significant associations with proteins. A couple of associations between proteins and mental health were even found with two different self-report measurements such as MPNI, CDI or GBI (e.g., Osteomodulin and Cadherin-13), while a couple of protein associations were found with adolescent ratings of both self-report and co-twin reports (e.g., IgGFc-binding protein and Complement C1q subcomponent subunit C). The effectiveness of adolescent self-reports to identify associations between mental health and metabolites^14,15^ or future psychiatric disorder^44^ has previously been seen in our earlier studies, and others have also found adolescent self reports or ratings from other youth (e.g., peers) to also be important and unique characterizations of mental health, possibly because they offer a view across environments such as home, school and leisure activities^45,46^. However, some studies, for example on ADHD, have not seen this same pattern^47^. In general though, it seems that adolescent self-reports for metabolomic and proteomic associations should be considered potentially valuable, and used at least along with other raters in future studies. While reports from parents or teachers may accurately reflect external perceptions of depressive symptoms or aggressive behavior, self-reports may be more reflective of the individual’s overall health. Additional investigations focusing on differences in proteomic associations across raters and different contexts may pave the way to a better understanding of protein functions in dimensions of mental health.

This study has many strengths including using multiple omics levels, mental health indicators, and raters of mental health, however, it is also important to consider some limitations. For example, our proteomics panel (and metabolomics panel) only included a portion of the entire proteome (and metabolome), thus as panels with a higher number of proteins become available, extending these analyses would be important. Additionally, despite not looking at the entire proteome, correcting for multiple testing limited our ability to identify associations and interactions. Another limitation is that we did not examine the role of medication use or substance use in confounding associations between plasma proteins and mental health indicators. A recent large-scale study identified proteins associated with medication use and smoking, suggesting that these variables influence the proteome^13^. In the current study, we found no significant association between depression medication use reported at age at blood collection and plasma proteins (number of cases: 20 out of 730), suggesting that depression medication use may play a minor role in confounding the reported associations. Lastly, since we only had proteomic and metabolomic data available at one time point (age 22), we are unable to draw any causal conclusions. Additionally, we used both mental health indicators at the same time point as the blood draw as well as earlier, not to suggest causal pathways, but to indicate whether patterns of associations were consistent, as we have seen in our previous investigations^14,15^, thus helping to indicate which biological pathways might be useful to investigate first. For causal pathways to be investigated, one could use Mendelian randomization^48^ or collect and investigate longitudinal changes in mental health and omics data. Longitudinal data could, among other things, help us to better understand the omic profiles of those with persistent poor mental health and/or those proteins that are sensitive to fluctuations in mental health over short- or mid-term time periods.

In conclusion, this study investigated proteomic associations between depressive symptoms and the p-factor using multiple mental health indicators and raters, as well as being first to identify a protein–metabolite interaction between two proteins (Mast/stem cell growth factor receptor Kit and Contactin-1) and valine (metabolite) in relation to p-factor levels. Most proteins were associated with the p-factor or both the p-factor and depressive symptoms, and several of the significant proteins have been associated with functions and disorders in the brain or are involved in immunity or inflammation, for example. This adds to our newly emerging and growing list of possible biological molecules and pathways to investigate for improving our understanding of the etiology, severity, and treatment options in mental health disorders.

## Methods

### Cohort and participants

The FinnTwin12 cohort was used in the following investigation. FinnTwin12 is a Finnish population-based cohort of twins born 1983–1987 with data collected from parents, teachers, classmates, and twins themselves at ages 11/12, 14, 17, and approximately 22 years old^49^. Data collection involved several questionnaires over the different waves of collection, and, at age 14 and 22, a more intensively studied subset of twins underwent semi-structured psychiatric interviews, additional questionnaires, and had biological samples collected.

Only individuals for whom plasma proteomics were obtained (from the intensive subset) were selected for the present analysis. Pregnant women (n=53) and those taking cholesterol medication (n=1) were also excluded, leading to a final sample size of N=730 twins. This sample comprised 56% females, and participants were 22.3 years old on average at the time of blood sampling (sd: 0.6; range: 21.0-25.0).

For each of the first three data collection waves, BMI was calculated from self-reported height and weight. In the wave corresponding to age 22 years, we calculated the BMI from the height and weight measurements that were clinically assessed on the day of the visit for the blood sample. Missing BMI information for each wave did not exceed 10% (range: 0-9.9%) and represented 4.8% of all BMI measures available. Missing BMI information was therefore imputed using the median BMI of each wave. Mean BMI was 17.6 (sd=2.6), 19.4 (sd=2.7), 21.4 (sd=2.7) and 23.3 (sd=4.0) kg.m^−2^ at ages 12, 14, 17 and 22, respectively.

Ethical approval for all data collection waves was obtained from the ethical committee of the Helsinki and Uusimaa University Hospital District and Indiana University’s Institutional Review Board. At ages 12 and 14, parents of the twins provided consent for the twins’ participation, while the twins themselves provided written consent at ages 17 and 22.

### Questionnaires and indicators of mental health

At age 12, 14, and 17 data collection waves, different raters were asked to fill out the modified Multidimensional Peer Nomination Inventory (MPNI) questions regarding the twins^50,51^. Raters included parents and teachers at age 12, teachers, the twins themselves and their co-twin at age 14, and the twins themselves and their co-twin at age 17. The MPNI can be used at multiple subscale and dimensional levels, and captures general levels of emotional and behavioral problems. Subscales include depression and social anxiety (these two are part of the internalizing dimensional scale), and aggressive behavior, hyperactivity/impulsivity, and inattention (these three comprise the externalizing dimensional scale). Additionally, the internalizing and externalizing problem dimensions can be summarized into a p-factor scale (thought disorder symptoms were not collected)^14^. A mean score is used, allowing no missing values for subscales and 4 missing values for the p-factor scale (missing items imputed to the mean score). Additionally, a “combined” p-factor score was created using the p-factor scores of all 7 of the available MPNI ratings (Cronbach’s alpha=0.76), because we know that ratings from different raters are not systematically, highly correlated^52,15^, even though factor analysis suggested a unique factor to be retained. For this combined p-factor, we therefore averaged the 7 individual p-factors so as to capture shared variability across different p-factors, which we previously scaled^12^ to mean zero and variance one.

At the age 14 and 22 data collection waves, semi-structured psychiatric interviews (Semi-Structured Assessment for the Genetics of Alcoholism; SSAGA^53^) occurred which included DSM-IV (Diagnostic and Statistical Manual of Mental Disorders) criteria that can be used to create psychiatric diagnoses, including MDD. For the assessment at ages 14, we used the number of lifetime depressive symptoms^54^ and for age 22 MDD diagnosis (Table 1). The choice of using lifetime depressive symptoms rather than MDD diagnosis at age 14 was motivated by the fact that only 3 participants were diagnosed with MDD, too few for statistical modeling, whereas 120 participants reported having one or more depressive symptoms during their lifetime. In contrast, 103 participants were diagnosed with MDD at age 22. In the SSAGA, medication use (including anti-depressant medication use) data was also collected.

Additionally, at age 17 and 22 data collection waves, the twins filled out the General Behavior Inventory (GBI) questionnaire^55,56^. This self-rated modified depression questionnaire includes a validated 10 items (the original GBI has 73 items with multiple subscales). A sum score is used, with one missing value allowed.

Lastly, at the age 14 data collection wave, twins filled out the Children’s Depression Inventory (CDI^57^), a 27-item inventory assessing depressive symptoms in youth. Each question was associated with 3 responses that reflected the severity of the symptoms associated with depression and were coded as 0, 1, or 2. The CDI score was defined as the sum of these scores, thus ranging from 0 to 54, with higher scores referring to higher levels of depressive symptoms. A maximum of 2 missing responses was allowed (59 and 19 participants had 1 or 2 missing items, respectively), and missing values were imputed with zeros, i.e., we did not increase the CDI score when missing values occurred, because missing values were not guaranteed to be randomly distributed.

Thus, there were 21 indicators of mental health used in total: MPNI parent age 12 depression (MPNI-p12d), MPNI teacher age 12 depression (MPNI-t12d), MPNI-t14d, MPNI self rating age 14 depression (MPNI-s14d), MPNI co-twin rating age 14 depression (MPNI-co14d), MPNI-s17d, MPNI-co17d, MPNI parent age 12 p-factor (MPNI-p12p), MPNI-t12p, MPNI-t14p, MPNI-s14p, MPNI-co14p, MPNI-s17p, MPNI-co17p, MPNI p-factor summary z-score (MPNI-pz), MDD age 14 (symptom count; MDD-14), MDD-22 (yes/no), depression medication use age 22 (yes/no; medication-22d), GBI age 17 (GBI-17), GBI-22, and CDI age 14 (CDI-14). Descriptive summaries of the 21 mental health indicators regarding Ns, means, standard deviations, ranges, skewness, and kurtosis can be found in Table 1. Additionally, correlations between continuous mental health indicators are shown in Figure 2A-B as both a network (A: abs(cor)>0.2) or correlation matrix (B).

### Omics processing

At the wave corresponding to age 22 years, blood plasma samples were collected after overnight fasting, with the request to have abstained from alcohol and tobacco since the night before sampling. In 2010, the samples were processed to obtain metabolomics data at Nightingale (formerly, Brainshake). In 2022, the samples were processed to obtain proteomics data at the Turku Proteomics Facility (Turku Proteomics Facility, Turku, Finland). Further details of proteomic and metabolomic data processing are described in the corresponding subsections.

#### Proteomic data processing

Proteins from plasma samples of the 730 individuals used in this study were processed in four batches at the Turku Proteomics Facility (Turku Proteomics Facility, Turku, Finland) using their LC-ESI-MS/MS (Q Exactive HF mass spectrometer) proteomics platform; a detailed description of the data is provided elsewhere^12^. Proteins were subjected to precipitation and in-solution digestion in accordance with the standard protocol of the Turku Proteomics Facility. A commercial kit (High Select™ Top14 Abundant Protein Depletion Mini Spin Columns, cat. No: A36370, ThermoScientific) was used to deplete the most abundant proteins from plasma prior to proteomic analysis. The data were first analyzed using Spectronaut software and included local normalization of the data^58^. Raw data was processed as described in detail elsewhere^10^. Briefly, data processing included log_2_ transformation of protein values, assessment of outliers, exclusion of proteins with >10% missing values, imputation of missing values using the lowest observed value per batch, and corrections for batch effects using Combat^59^. The protein abundances were scaled such that one unit corresponded to one standard deviation with a mean of zero. The final proteomic dataset consisted of 439 proteins for the 730 individuals in the study.

#### Metabolomic data processing

Metabolites from plasma samples of the 730 individuals used in this study were processed in one batch at Nightingale (formerly, Brainshake) using their automated high-throughput ^1^H nuclear magnetic resonance spectroscopy (NMR) metabolomics platform^14,15^. Metabolite values were available in mmol/l, and were log transformed. We focused on three specific metabolites in this study that had already been associated in FinnTwin12 with mental health: 3-hydroxybutyrate was negatively associated with aggressive behavior and the p-factor^14,15^ and valine and leucine were negatively associated with depression^14^.

### Statistical analyses

We first quantified the associations between protein levels and mental health indicators using linear and generalized mixed-effects models. We successively modeled each mental health indicator (n=21) as a dependent variable while proteins were used as independent variables. Age at blood sampling, sex and BMI at blood sampling were used as covariates. In models where mental health indicators were derived from questionnaires completed at ages 12, 14 or 17, BMI at 12, 14 or 17 years of age, respectively, was also included as a covariate. The inclusion of adolescent BMI was performed because the variability of BMI at blood sampling was not found to be strongly determined by self-reported BMI during adolescence, since the coefficients of determination (R-square) derived from univariate linear models were in the range of 37-52%. This ensured identification of proteins associated with adolescent mental health independent of both adolescent and adult BMI. For models using the z-scored p-factor, given that the p-factor was averaged across different ages, we did not include BMI measurements assessed during adolescence but only BMI at blood sampling. To correct for family relatedness in the data, we used family identifiers as random effects. Depending on whether the mental health indicator assessed was a continuous or binary outcome, we used linear and generalized mixed-effects models, respectively. Nullity of fixed-effect coefficients related to proteins was tested, from which nominal p-values were derived. For each set of associations between a mental health indicator and the 439 proteins, we used the Benjamini– Hochberg procedure to control the False Discovery Rate (FDR; α=0.20).

In a follow-up analysis (Figure 1), we sought to determine whether, when significant associations between protein levels and mental health indicators were identified, the addition of interaction terms with metabolites reported in the literature could deepen our understanding of the molecular basis of mental health. We carried out interaction analyses by adding fixed-effect coefficients for the metabolite and its interaction with the protein to the initial model(s) described above. Due to limited statistical power to test for interactions between proteins and metabolites, we restricted interaction analyses to proteins that were strongly associated with mental health indicators, i.e., with a standardized coefficient above 3 in absolute value (Figure 1). In addition, interaction analyses were performed if a metabolite was found to be sufficiently associated with the mental health indicator to be modeled (nominal p<0.05), as assessed by initial linear or generalized mixed effects models described above. Interaction nullity was assessed by t- or z-test, and interactions were considered significant if nominal p-values were below 0.05. Models were run using the R package lme4 version 1.1-30 under the R Studio environment (version 4.1.3).

## Data access

The data used in the analysis is deposited in the Biobank of the Finnish Institute for Health and Welfare (https://thl.fi/en/web/thl-biobank/forresearchers). It is available to researchers after written application and following the relevant Finnish legislation.

## Authors’ contributions

AMW prepared the dataset, drafted the research plan, assisted with the data analysis, co-drafted the first draft of the manuscript, helped edit the manuscript; GD contributed to the research plan, ran the data analyses, created the Tables and Figures, co-drafted the first draft of the manuscript, helped edit the manuscript; RJR secured funding for the data collection, helped establish the cohort, gave critical feedback on the manuscript, helped in editing the manuscript; LP helped establish the cohort, gave critical feedback on the manuscript; JK secured funding for the data collection, established the cohort, contributed to the research plan, assisted with interpretation of results, gave critical feedback on the manuscript

---

Table S1: Associations between protein abundance and mental health indicators with nominal p-value under 0.05.

**Legend:** Associations between protein levels and mental health indicators are shown regardless of any multiple correction (nominal p-value below 0.05). FDR: False Discovery Rate. se: standard error.

