## Supplemental Table 1 for "Protein associations and protein–metabolite interactions with depressive symptoms and the p-factor"

| | | Protein description | | Fixed-effect coefficient | | | P-value | | |
|---|---|---|---|---|---|---|---|---|---|
| Indicator | Protein | Genes | Description | Estimate | se | t/z.values | nominal | FDR | Bonferroni |
| CDI-14 | O15143 | ARPC1B | Actin-related protein 2/3 complex subunit 1B | -0.43 | 0.2 | -2.3 | 2.0E-02 | 7.8E-01 | 1.0E+00 |
| CDI-14 | O75144 | ICOSLG | ICOS ligand | -0.40 | 0.2 | -2.2 | 2.9E-02 | 7.8E-01 | 1.0E+00 |
| CDI-14 | O94919 | ENDOD1 | Endonuclease domain-containing 1 protein | 0.37 | 0.2 | 2.0 | 4.6E-02 | 9.1E-01 | 1.0E+00 |
| CDI-14 | P00740 | F9 | Coagulation factor IX | 0.39 | 0.2 | 2.1 | 3.6E-02 | 8.3E-01 | 1.0E+00 |
| CDI-14 | P00751 | CFB | Complement factor B | 0.45 | 0.2 | 2.4 | 1.7E-02 | 7.8E-01 | 1.0E+00 |
| CDI-14 | P02743 | APCS | Serum amyloid P-component | 0.46 | 0.2 | 2.2 | 2.9E-02 | 7.8E-01 | 1.0E+00 |
| CDI-14 | P04746 | AMY2A | Pancreatic alpha-amylase | -0.40 | 0.2 | -2.2 | 2.9E-02 | 7.8E-01 | 1.0E+00 |
| CDI-14 | P08253 | MMP2 | 72 kDa type IV collagenase | -0.42 | 0.2 | -2.2 | 3.0E-02 | 7.8E-01 | 1.0E+00 |
| CDI-14 | P08294 | SOD3 | Extracellular superoxide dismutase [Cu-Zn] | 0.43 | 0.2 | 2.1 | 3.5E-02 | 8.3E-01 | 1.0E+00 |
| CDI-14 | P09960 | LTA4H | Leukotriene A-4 hydrolase | 0.36 | 0.2 | 2.0 | 4.4E-02 | 9.1E-01 | 1.0E+00 |
| CDI-14 | P0C0L5 | C4B | Complement C4-B | 0.54 | 0.2 | 2.8 | 5.4E-03 | 4.7E-01 | 1.0E+00 |
| CDI-14 | P0DJI9 | SAA2 | Serum amyloid A-2 protein | -0.44 | 0.2 | -2.3 | 2.4E-02 | 7.8E-01 | 1.0E+00 |
| CDI-14 | P11362 | FGFR1 | Fibroblast growth factor receptor 1 | -0.42 | 0.2 | -2.3 | 2.2E-02 | 7.8E-01 | 1.0E+00 |
| CDI-14 | P43121 | MCAM | Cell surface glycoprotein MUC18 | -0.44 | 0.2 | -2.3 | 2.2E-02 | 7.8E-01 | 1.0E+00 |
| CDI-14 | P55103 | INHBC | Inhibin beta C chain | 0.54 | 0.2 | 3.0 | 2.9E-03 | 4.7E-01 | 1.0E+00 |
| CDI-14 | P55290 | CDH13 | Cadherin-13 | -0.50 | 0.2 | -2.6 | 1.1E-02 | 6.6E-01 | 1.0E+00 |
| CDI-14 | P69892 | HBG2 | Hemoglobin subunit gamma-2 | -0.48 | 0.2 | -2.7 | 8.1E-03 | 6.0E-01 | 1.0E+00 |
| CDI-14 | Q12860 | CNTN1 | Contactin-1 | -0.42 | 0.2 | -2.2 | 2.9E-02 | 7.8E-01 | 1.0E+00 |
| CDI-14 | Q13449 | LSAMP | Limbic system-associated membrane protein | -0.57 | 0.2 | -2.9 | 3.3E-03 | 4.7E-01 | 1.0E+00 |
| CDI-14 | Q99983 | OMD | Osteomodulin | -0.65 | 0.2 | -3.6 | 3.4E-04 | 1.5E-01 | 1.5E-01 |
| CDI-14 | Q9NY15 | STAB1 | Stabilin-1 | -0.52 | 0.2 | -2.8 | 4.9E-03 | 4.7E-01 | 1.0E+00 |
| MDD-14 | O14791 | APOL1 | Apolipoprotein L1 | 0.05 | 0.0 | 2.1 | 4.0E-02 | 8.5E-01 | 1.0E+00 |
| MDD-14 | P00918 | CA2 | Carbonic anhydrase 2 | -0.05 | 0.0 | -2.2 | 2.6E-02 | 8.5E-01 | 1.0E+00 |
| MDD-14 | P01024 | C3 | Complement C3 | 0.05 | 0.0 | 2.0 | 4.5E-02 | 8.5E-01 | 1.0E+00 |
| MDD-14 | P01137 | TGFB1 | Transforming growth factor beta-1 proprotein | 0.05 | 0.0 | 2.0 | 4.3E-02 | 8.5E-01 | 1.0E+00 |
| MDD-14 | P01344 | IGF2 | Insulin-like growth factor II | -0.05 | 0.0 | -2.0 | 4.8E-02 | 8.5E-01 | 1.0E+00 |
| MDD-14 | P02533 | KRT14 | Keratin, type I cytoskeletal 14 | 0.05 | 0.0 | 2.0 | 4.3E-02 | 8.5E-01 | 1.0E+00 |
| MDD-14 | P02741 | CRP | C-reactive protein | 0.05 | 0.0 | 2.0 | 4.3E-02 | 8.5E-01 | 1.0E+00 |
| MDD-14 | P03950 | ANG | Angiogenin | -0.05 | 0.0 | -2.1 | 3.3E-02 | 8.5E-01 | 1.0E+00 |
| MDD-14 | P05156 | CFI | Complement factor I | 0.06 | 0.0 | 2.5 | 1.4E-02 | 8.5E-01 | 1.0E+00 |
| MDD-14 | P05160 | F13B | Coagulation factor XIII B chain | 0.06 | 0.0 | 2.5 | 1.2E-02 | 8.5E-01 | 1.0E+00 |

| | | | | | | | | | |
|---|---|---|---|---|---|---|---|---|---|
| MDD-14 | P05543 | SERPINA7 | Thyroxine-binding globulin | 0.05 | 0.0 | 2.1 | 3.9E-02 | 8.5E-01 | 1.0E+00 |
| MDD-14 | P07307 | ASGR2 | Asialoglycoprotein receptor 2 | -0.04 | 0.0 | -2.0 | 4.8E-02 | 8.5E-01 | 1.0E+00 |
| MDD-14 | P17936 | IGFBP3 | Insulin-like growth factor-binding protein 3 | -0.06 | 0.0 | -2.5 | 1.4E-02 | 8.5E-01 | 1.0E+00 |
| MDD-14 | P18065 | IGFBP2 | Insulin-like growth factor-binding protein 2 | -0.05 | 0.0 | -2.0 | 4.4E-02 | 8.5E-01 | 1.0E+00 |
| MDD-14 | P33151 | CDH5 | Cadherin-5 | -0.07 | 0.0 | -3.1 | 2.0E-03 | 8.5E-01 | 8.7E-01 |
| MDD-14 | P35527 | KRT9 | Keratin, type I cytoskeletal 9 | 0.05 | 0.0 | 2.1 | 3.8E-02 | 8.5E-01 | 1.0E+00 |
| MDD-14 | P36980 | CFHR2 | Complement factor H-related protein 2 | 0.05 | 0.0 | 2.0 | 4.2E-02 | 8.5E-01 | 1.0E+00 |
| MDD-14 | P40189 | IL6ST | Interleukin-6 receptor subunit beta | -0.06 | 0.0 | -2.7 | 7.2E-03 | 8.5E-01 | 1.0E+00 |
| MDD-14 | P80723 | BASP1 | Brain acid soluble protein 1 | -0.04 | 0.0 | -2.0 | 4.8E-02 | 8.5E-01 | 1.0E+00 |
| MDD-14 | P98160 | HSPG2 | Basement membrane-specific heparan sulfate proteoglycan core protein | -0.06 | 0.0 | -2.5 | 1.1E-02 | 8.5E-01 | 1.0E+00 |
| MDD-14 | Q01459 | CTBS | Di-N-acetylchitobiase | 0.05 | 0.0 | 2.0 | 4.4E-02 | 8.5E-01 | 1.0E+00 |
| MDD-14 | Q9BXR6 | CFHR5 | Complement factor H-related protein 5 | 0.05 | 0.0 | 2.1 | 3.4E-02 | 8.5E-01 | 1.0E+00 |
| MDD-14 | Q9H1U4 | MEGF9 | Multiple epidermal growth factor-like domains protein 9 | 0.05 | 0.0 | 2.3 | 2.2E-02 | 8.5E-01 | 1.0E+00 |
| MDD-14 | Q9UBX1 | CTSF | Cathepsin F | -0.04 | 0.0 | -2.0 | 4.8E-02 | 8.5E-01 | 1.0E+00 |
| GBI-17 | P03951 | F11 | Coagulation factor XI | -0.39 | 0.2 | -2.1 | 4.0E-02 | 1.0E+00 | 1.0E+00 |
| GBI-17 | P04264 | KRT1 | Keratin, type II cytoskeletal 1 | -0.36 | 0.2 | -2.0 | 4.6E-02 | 1.0E+00 | 1.0E+00 |
| GBI-17 | P05062 | ALDOB | Fructose-bisphosphate aldolase B | 0.41 | 0.2 | 2.1 | 3.5E-02 | 1.0E+00 | 1.0E+00 |
| GBI-17 | P06702 | S100A9 | Protein S100-A9 | -0.37 | 0.2 | -2.0 | 4.7E-02 | 1.0E+00 | 1.0E+00 |
| GBI-17 | P08603 | CFH | Complement factor H | -0.39 | 0.2 | -2.0 | 4.6E-02 | 1.0E+00 | 1.0E+00 |
| GBI-17 | P10586 | PTPRF | Receptor-type tyrosine-protein phosphatase F | -0.60 | 0.2 | -3.4 | 7.1E-04 | 3.1E-01 | 3.1E-01 |
| GBI-17 | P11717 | IGF2R | Cation-independent mannose-6-phosphate receptor | -0.37 | 0.2 | -2.1 | 4.1E-02 | 1.0E+00 | 1.0E+00 |
| GBI-17 | P24593 | IGFBP5 | Insulin-like growth factor-binding protein 5 | -0.44 | 0.2 | -2.3 | 2.3E-02 | 1.0E+00 | 1.0E+00 |
| GBI-17 | P30043 | BLVRB | Flavin reductase (NADPH) | -0.38 | 0.2 | -2.1 | 4.0E-02 | 1.0E+00 | 1.0E+00 |
| GBI-17 | P32119 | PRDX2 | Peroxiredoxin-2 | -0.43 | 0.2 | -2.4 | 1.9E-02 | 1.0E+00 | 1.0E+00 |
| GBI-17 | P48594 | SERPINB4 | Serpin B4 | -0.37 | 0.2 | -2.1 | 3.7E-02 | 1.0E+00 | 1.0E+00 |
| GBI-17 | P61626 | LYZ | Lysozyme C | 0.38 | 0.2 | 2.0 | 4.7E-02 | 1.0E+00 | 1.0E+00 |
| GBI-17 | Q13449 | LSAMP | Limbic system-associated membrane protein | -0.39 | 0.2 | -2.0 | 4.1E-02 | 1.0E+00 | 1.0E+00 |
| GBI-17 | Q15063 | POSTN | Periostin | -0.40 | 0.2 | -2.2 | 3.2E-02 | 1.0E+00 | 1.0E+00 |
| GBI-17 | Q15113 | PCOLCE | Procollagen C-endopeptidase enhancer 1 | 0.40 | 0.2 | 2.1 | 3.3E-02 | 1.0E+00 | 1.0E+00 |
| GBI-17 | Q562R1 | ACTBL2 | Beta-actin-like protein 2 | 0.36 | 0.2 | 2.0 | 4.4E-02 | 1.0E+00 | 1.0E+00 |

| | | | | | | | | | |
|---|---|---|---|---|---|---|---|---|---|
| GBI-17 | Q6YHK3 | CD109 | CD109 antigen | -0.38 | 0.2 | -2.0 | 4.5E-02 | 1.0E+00 | 1.0E+00 |
| GBI-17 | Q8N1F8 | STK11IP | Serine/threonine-protein kinase 11-interacting protein | 0.39 | 0.2 | 2.1 | 3.3E-02 | 1.0E+00 | 1.0E+00 |
| GBI-17 | Q99983 | OMD | Osteomodulin | -0.42 | 0.2 | -2.2 | 2.6E-02 | 1.0E+00 | 1.0E+00 |
| GBI-22 | A1L4H1 | SSC5D | Soluble scavenger receptor cysteine-rich domain-containing protein SSC5D | 0.37 | 0.2 | 2.1 | 3.2E-02 | 4.4E-01 | 1.0E+00 |
| GBI-22 | O00533 | CHL1 | Neural cell adhesion molecule L1-like protein | -0.36 | 0.2 | -2.0 | 4.4E-02 | 4.6E-01 | 1.0E+00 |
| GBI-22 | O75594 | PGLYRP1 | Peptidoglycan recognition protein 1 | 0.35 | 0.2 | 2.2 | 3.2E-02 | 4.4E-01 | 1.0E+00 |
| GBI-22 | P00488 | F13A1 | Coagulation factor XIII A chain | 0.34 | 0.2 | 2.0 | 4.3E-02 | 4.6E-01 | 1.0E+00 |
| GBI-22 | P00747 | PLG | Plasminogen | -0.39 | 0.2 | -2.1 | 3.3E-02 | 4.4E-01 | 1.0E+00 |
| GBI-22 | P02649 | APOE | Apolipoprotein E | 0.44 | 0.2 | 2.5 | 1.2E-02 | 4.4E-01 | 1.0E+00 |
| GBI-22 | P02655 | APOC2 | Apolipoprotein C-II | 0.39 | 0.2 | 2.2 | 2.9E-02 | 4.4E-01 | 1.0E+00 |
| GBI-22 | P02750 | LRG1 | Leucine-rich alpha-2-glycoprotein | 0.41 | 0.2 | 2.3 | 2.5E-02 | 4.4E-01 | 1.0E+00 |
| GBI-22 | P02760 | AMBP | Protein AMBP | -0.35 | 0.2 | -2.1 | 4.0E-02 | 4.6E-01 | 1.0E+00 |
| GBI-22 | P02766 | TTR | Transthyretin | -0.47 | 0.2 | -2.5 | 1.2E-02 | 4.4E-01 | 1.0E+00 |
| GBI-22 | P04004 | VTN | Vitronectin | 0.39 | 0.2 | 2.1 | 3.7E-02 | 4.4E-01 | 1.0E+00 |
| GBI-22 | P05062 | ALDOB | Fructose-bisphosphate aldolase B | 0.41 | 0.2 | 2.4 | 1.6E-02 | 4.4E-01 | 1.0E+00 |
| GBI-22 | P05121 | SERPINE1 | Plasminogen activator inhibitor 1 | 0.36 | 0.2 | 2.2 | 3.1E-02 | 4.4E-01 | 1.0E+00 |
| GBI-22 | P05164 | MPO | Myeloperoxidase | 0.45 | 0.2 | 2.7 | 7.7E-03 | 4.4E-01 | 1.0E+00 |
| GBI-22 | P06396 | GSN | Gelsolin | -0.40 | 0.2 | -2.2 | 2.7E-02 | 4.4E-01 | 1.0E+00 |
| GBI-22 | P06681 | C2 | Complement C2 | 0.37 | 0.2 | 2.2 | 2.8E-02 | 4.4E-01 | 1.0E+00 |
| GBI-22 | P07339 | CTSD | Cathepsin D | 0.38 | 0.2 | 2.3 | 2.4E-02 | 4.4E-01 | 1.0E+00 |
| GBI-22 | P07942 | LAMB1 | Laminin subunit beta-1 | -0.37 | 0.2 | -2.2 | 2.7E-02 | 4.4E-01 | 1.0E+00 |
| GBI-22 | P07996 | THBS1 | Thrombospondin-1 | 0.37 | 0.2 | 2.1 | 3.3E-02 | 4.4E-01 | 1.0E+00 |
| GBI-22 | P11362 | FGFR1 | Fibroblast growth factor receptor 1 | -0.40 | 0.2 | -2.5 | 1.4E-02 | 4.4E-01 | 1.0E+00 |
| GBI-22 | P11597 | CETP | Cholesteryl ester transfer protein | -0.42 | 0.2 | -2.5 | 1.4E-02 | 4.4E-01 | 1.0E+00 |
| GBI-22 | P14780 | MMP9 | Matrix metalloproteinase-9 | 0.34 | 0.2 | 2.0 | 4.4E-02 | 4.6E-01 | 1.0E+00 |
| GBI-22 | P16112 | ACAN | Aggrecan core protein | -0.36 | 0.2 | -2.1 | 3.3E-02 | 4.4E-01 | 1.0E+00 |
| GBI-22 | P24043 | LAMA2 | Laminin subunit alpha-2 | -0.39 | 0.2 | -2.3 | 2.1E-02 | 4.4E-01 | 1.0E+00 |
| GBI-22 | P27169 | PON1 | Serum paraoxonase/arylesterase 1 | -0.36 | 0.2 | -2.1 | 3.6E-02 | 4.4E-01 | 1.0E+00 |
| GBI-22 | P32119 | PRDX2 | Peroxiredoxin-2 | -0.37 | 0.2 | -2.3 | 2.2E-02 | 4.4E-01 | 1.0E+00 |
| GBI-22 | P33151 | CDH5 | Cadherin-5 | -0.57 | 0.2 | -3.2 | 1.3E-03 | 2.0E-01 | 5.5E-01 |
| GBI-22 | P35542 | SAA4 | Serum amyloid A-4 protein | 0.36 | 0.2 | 2.1 | 4.0E-02 | 4.6E-01 | 1.0E+00 |
| GBI-22 | P49913 | CAMP | Cathelicidin antimicrobial peptide | 0.37 | 0.2 | 2.2 | 2.9E-02 | 4.4E-01 | 1.0E+00 |
| GBI-22 | P55290 | CDH13 | Cadherin-13 | -0.56 | 0.2 | -3.2 | 1.4E-03 | 2.0E-01 | 6.0E-01 |
| GBI-22 | P61626 | LYZ | Lysozyme C | 0.46 | 0.2 | 2.7 | 7.7E-03 | 4.4E-01 | 1.0E+00 |
| GBI-22 | P61769 | B2M | Beta-2-microglobulin | 0.36 | 0.2 | 2.1 | 3.5E-02 | 4.4E-01 | 1.0E+00 |
| GBI-22 | P69892 | HBG2 | Hemoglobin subunit gamma-2 | -0.39 | 0.2 | -2.4 | 1.6E-02 | 4.4E-01 | 1.0E+00 |

| | | | | | | | | | |
|---|---|---|---|---|---|---|---|---|---|
| GBI-22 | Q12841 | FSTL1 | Follistatin-related protein 1 | 0.37 | 0.2 | 2.2 | 3.1E-02 | 4.4E-01 | 1.0E+00 |
| GBI-22 | Q12860 | CNTN1 | Contactin-1 | -0.40 | 0.2 | -2.4 | 1.8E-02 | 4.4E-01 | 1.0E+00 |
| GBI-22 | Q13201 | MMRN1 | Multimerin-1 | 0.45 | 0.2 | 2.7 | 8.1E-03 | 4.4E-01 | 1.0E+00 |
| GBI-22 | Q6UXB8 | PI16 | Peptidase inhibitor 16 | -0.35 | 0.2 | -2.1 | 3.7E-02 | 4.4E-01 | 1.0E+00 |
| GBI-22 | Q86SQ4 | ADGRG6 | Adhesion G-protein coupled receptor G6 | 0.41 | 0.2 | 2.3 | 2.5E-02 | 4.4E-01 | 1.0E+00 |
| GBI-22 | Q86VB7 | CD163 | Scavenger receptor cysteine-rich type 1 protein M130 | 0.58 | 0.2 | 3.3 | 9.1E-04 | 2.0E-01 | 4.0E-01 |
| GBI-22 | Q92954 | PRG4 | Proteoglycan 4 | 0.61 | 0.2 | 3.1 | 1.8E-03 | 2.0E-01 | 7.9E-01 |
| GBI-22 | Q99983 | OMD | Osteomodulin | -0.42 | 0.2 | -2.6 | 1.0E-02 | 4.4E-01 | 1.0E+00 |
| GBI-22 | Q9Y6Z7 | COLEC10 | Collectin-10 | -0.39 | 0.2 | -2.3 | 2.1E-02 | 4.4E-01 | 1.0E+00 |
| MDD-22 | P00746 | CFD | Complement factor D | 0.31 | 0.1 | 2.4 | 1.6E-02 | 8.3E-01 | 1.0E+00 |
| MDD-22 | P01034 | CST3 | Cystatin-C | 0.26 | 0.1 | 2.1 | 3.7E-02 | 8.3E-01 | 1.0E+00 |
| MDD-22 | P02745 | C1QA | Complement C1q subcomponent subunit A | -0.26 | 0.1 | -2.3 | 2.0E-02 | 8.3E-01 | 1.0E+00 |
| MDD-22 | P02746 | C1QB | Complement C1q subcomponent subunit B | -0.38 | 0.1 | -3.1 | 1.7E-03 | 7.4E-01 | 7.4E-01 |
| MDD-22 | P03951 | F11 | Coagulation factor XI | -0.28 | 0.1 | -2.4 | 1.6E-02 | 8.3E-01 | 1.0E+00 |
| MDD-22 | P04075 | ALDOA | Fructose-bisphosphate aldolase A | -0.26 | 0.1 | -2.1 | 3.3E-02 | 8.3E-01 | 1.0E+00 |
| MDD-22 | P05155 | SERPING1 | Plasma protease C1 inhibitor | -0.25 | 0.1 | -2.2 | 3.1E-02 | 8.3E-01 | 1.0E+00 |
| MDD-22 | P05156 | CFI | Complement factor I | -0.33 | 0.1 | -2.9 | 4.2E-03 | 8.3E-01 | 1.0E+00 |
| MDD-22 | P08603 | CFH | Complement factor H | -0.25 | 0.1 | -2.1 | 3.8E-02 | 8.3E-01 | 1.0E+00 |
| MDD-22 | P11047 | LAMC1 | Laminin subunit gamma-1 | -0.23 | 0.1 | -2.3 | 1.9E-02 | 8.3E-01 | 1.0E+00 |
| MDD-22 | P12109 | COL6A1 | Collagen alpha-1(VI) chain | -0.24 | 0.1 | -2.2 | 3.0E-02 | 8.3E-01 | 1.0E+00 |
| MDD-22 | P13473 | LAMP2 | Lysosome-associated membrane glycoprotein 2 | 0.24 | 0.1 | 2.1 | 3.8E-02 | 8.3E-01 | 1.0E+00 |
| MDD-22 | P13727 | PRG2 | Bone marrow proteoglycan | -0.24 | 0.1 | -2.1 | 3.7E-02 | 8.3E-01 | 1.0E+00 |
| MDD-22 | P14618 | PKM | Pyruvate kinase PKM | -0.32 | 0.1 | -2.6 | 9.8E-03 | 8.3E-01 | 1.0E+00 |
| MDD-22 | P24043 | LAMA2 | Laminin subunit alpha-2 | -0.22 | 0.1 | -2.4 | 1.9E-02 | 8.3E-01 | 1.0E+00 |
| MDD-22 | P34096 | RNASE4 | Ribonuclease 4 | 0.27 | 0.1 | 2.2 | 2.8E-02 | 8.3E-01 | 1.0E+00 |
| MDD-22 | P50395 | GDI2 | Rab GDP dissociation inhibitor beta | 0.28 | 0.1 | 2.2 | 2.8E-02 | 8.3E-01 | 1.0E+00 |
| MDD-22 | P60174 | TPI1 | Triosephosphate isomerase | -0.24 | 0.1 | -2.0 | 4.5E-02 | 8.3E-01 | 1.0E+00 |
| MDD-22 | P63261 | ACTG1 | Actin, cytoplasmic 2 | -0.28 | 0.1 | -2.4 | 1.6E-02 | 8.3E-01 | 1.0E+00 |
| MDD-22 | P68032.P68133 | NA | NA | -0.23 | 0.1 | -2.0 | 4.4E-02 | 8.3E-01 | 1.0E+00 |
| MDD-22 | Q12884 | FAP | Prolyl endopeptidase FAP | -0.24 | 0.1 | -2.2 | 3.0E-02 | 8.3E-01 | 1.0E+00 |
| MDD-22 | Q4LDE5 | SVEP1 | Sushi, von Willebrand factor type A, EGF and pentraxin domain-containing protein 1 | -0.23 | 0.1 | -2.2 | 2.8E-02 | 8.3E-01 | 1.0E+00 |
| MDD-22 | Q8NDA2 | HMCN2 | Hemicentin-2 | 0.24 | 0.1 | 2.0 | 4.1E-02 | 8.3E-01 | 1.0E+00 |

| | | | | | | | | | |
|---|---|---|---|---|---|---|---|---|---|
| MDD-22 | Q9Y6Z7 | COLEC10 | Collectin-10 | -0.23 | 0.1 | -2.0 | 4.3E-02 | 8.3E-01 | 1.0E+00 |
| dep_meds | O75882 | ATRN | Attractin | 0.49 | 0.2 | 2.0 | 4.5E-02 | 9.9E-01 | 1.0E+00 |
| dep_meds | P00338 | LDHA | L-lactate dehydrogenase A chain | -0.49 | 0.2 | -2.2 | 2.4E-02 | 9.9E-01 | 1.0E+00 |
| dep_meds | P00915 | CA1 | Carbonic anhydrase 1 | -0.41 | 0.2 | -2.4 | 1.7E-02 | 9.9E-01 | 1.0E+00 |
| dep_meds | P03951 | F11 | Coagulation factor XI | -0.46 | 0.2 | -2.0 | 4.5E-02 | 9.9E-01 | 1.0E+00 |
| dep_meds | P04275 | VWF | von Willebrand factor | -0.54 | 0.2 | -2.4 | 1.6E-02 | 9.9E-01 | 1.0E+00 |
| dep_meds | P07195 | LDHB | L-lactate dehydrogenase B chain | -0.56 | 0.2 | -2.5 | 1.3E-02 | 9.9E-01 | 1.0E+00 |
| dep_meds | P08709 | F7 | Coagulation factor VII | 0.59 | 0.2 | 2.6 | 1.0E-02 | 9.9E-01 | 1.0E+00 |
| dep_meds | P13796 | LCP1 | Plastin-2 | -0.73 | 0.2 | -3.5 | 5.0E-04 | 2.2E-01 | 2.2E-01 |
| dep_meds | P22352 | GPX3 | Glutathione peroxidase 3 | -0.58 | 0.2 | -2.4 | 1.6E-02 | 9.9E-01 | 1.0E+00 |
| dep_meds | P48668 | KRT6C | Keratin, type II cytoskeletal 6C | -0.38 | 0.2 | -2.3 | 2.1E-02 | 9.9E-01 | 1.0E+00 |
| dep_meds | P53634 | CTSC | Dipeptidyl peptidase 1 | 0.57 | 0.3 | 2.2 | 2.7E-02 | 9.9E-01 | 1.0E+00 |
| dep_meds | P61769 | B2M | Beta-2-microglobulin | 0.49 | 0.2 | 2.0 | 4.8E-02 | 9.9E-01 | 1.0E+00 |
| dep_meds | Q12841 | FSTL1 | Follistatin-related protein 1 | 0.70 | 0.3 | 2.3 | 2.1E-02 | 9.9E-01 | 1.0E+00 |
| dep_meds | Q9BY67 | CADM1 | Cell adhesion molecule 1 | 1.90 | 0.7 | 2.7 | 7.1E-03 | 9.9E-01 | 1.0E+00 |
| MPNI-pz | O00592 | PODXL | Podocalyxin | -0.07 | 0.0 | -2.0 | 4.9E-02 | 4.8E-01 | 1.0E+00 |
| MPNI-pz | O14786 | NRP1 | Neuropilin-1 | -0.07 | 0.0 | -2.1 | 3.4E-02 | 4.4E-01 | 1.0E+00 |
| MPNI-pz | O75882 | ATRN | Attractin | -0.09 | 0.0 | -2.4 | 1.5E-02 | 3.1E-01 | 1.0E+00 |
| MPNI-pz | O95497 | VNN1 | Pantetheinase | 0.08 | 0.0 | 2.1 | 3.4E-02 | 4.4E-01 | 1.0E+00 |
| MPNI-pz | P02533 | KRT14 | Keratin, type I cytoskeletal 14 | -0.06 | 0.0 | -2.0 | 4.5E-02 | 4.7E-01 | 1.0E+00 |
| MPNI-pz | P02652 | APOA2 | Apolipoprotein A-II | 0.07 | 0.0 | 2.1 | 3.7E-02 | 4.4E-01 | 1.0E+00 |
| MPNI-pz | P02747 | C1QC | Complement C1q subcomponent subunit C | -0.07 | 0.0 | -2.1 | 3.3E-02 | 4.4E-01 | 1.0E+00 |
| MPNI-pz | P02760 | AMBP | Protein AMBP | -0.07 | 0.0 | -2.1 | 3.9E-02 | 4.5E-01 | 1.0E+00 |
| MPNI-pz | P03950 | ANG | Angiogenin | 0.08 | 0.0 | 2.2 | 2.5E-02 | 3.8E-01 | 1.0E+00 |
| MPNI-pz | P04070 | PROC | Vitamin K-dependent protein C | 0.07 | 0.0 | 2.0 | 4.8E-02 | 4.7E-01 | 1.0E+00 |
| MPNI-pz | P05160 | F13B | Coagulation factor XIII B chain | 0.08 | 0.0 | 2.4 | 1.8E-02 | 3.4E-01 | 1.0E+00 |
| MPNI-pz | P05546 | SERPIND1 | Heparin cofactor 2 | 0.09 | 0.0 | 2.6 | 8.4E-03 | 2.6E-01 | 1.0E+00 |
| MPNI-pz | P07339 | CTSD | Cathepsin D | 0.09 | 0.0 | 2.7 | 8.0E-03 | 2.6E-01 | 1.0E+00 |
| MPNI-pz | P07911 | UMOD | Uromodulin | -0.11 | 0.0 | -3.3 | 9.3E-04 | 8.1E-02 | 4.1E-01 |
| MPNI-pz | P08238 | HSP90AB1 | Heat shock protein HSP 90-beta | -0.06 | 0.0 | -2.0 | 4.4E-02 | 4.7E-01 | 1.0E+00 |
| MPNI-pz | P08571 | CD14 | Monocyte differentiation antigen CD14 | 0.13 | 0.0 | 4.0 | 7.5E-05 | 1.6E-02 | 3.3E-02 |
| MPNI-pz | P09172 | DBH | Dopamine beta-hydroxylase | -0.13 | 0.0 | -3.6 | 3.9E-04 | 4.8E-02 | 1.7E-01 |
| MPNI-pz | P09960 | LTA4H | Leukotriene A-4 hydrolase | 0.08 | 0.0 | 2.5 | 1.3E-02 | 2.9E-01 | 1.0E+00 |
| MPNI-pz | P0C0L5 | C4B | Complement C4-B | 0.08 | 0.0 | 2.3 | 2.4E-02 | 3.8E-01 | 1.0E+00 |
| MPNI-pz | P11047 | LAMC1 | Laminin subunit gamma-1 | -0.07 | 0.0 | -2.1 | 3.5E-02 | 4.4E-01 | 1.0E+00 |
| MPNI-pz | P12821 | ACE | Angiotensin-converting enzyme | -0.07 | 0.0 | -2.0 | 4.7E-02 | 4.7E-01 | 1.0E+00 |
| MPNI-pz | P14618 | PKM | Pyruvate kinase PKM | -0.09 | 0.0 | -2.5 | 1.3E-02 | 2.9E-01 | 1.0E+00 |
| MPNI-pz | P15169 | CPN1 | Carboxypeptidase N catalytic chain | -0.08 | 0.0 | -2.1 | 3.7E-02 | 4.4E-01 | 1.0E+00 |

| | | | | | | | | | |
|---|---|---|---|---|---|---|---|---|---|
| MPNI-pz | P19021 | PAM | Peptidyl-glycine alpha-amidating monooxygenase | 0.08 | 0.0 | 2.4 | 1.8E-02 | 3.4E-01 | 1.0E+00 |
| MPNI-pz | P19320 | VCAM1 | Vascular cell adhesion protein 1 | -0.08 | 0.0 | -2.3 | 2.1E-02 | 3.6E-01 | 1.0E+00 |
| MPNI-pz | P19827 | ITIH1 | Inter-alpha-trypsin inhibitor heavy chain H1 | -0.09 | 0.0 | -2.6 | 9.9E-03 | 2.9E-01 | 1.0E+00 |
| MPNI-pz | P23142 | FBLN1 | Fibulin-1 | -0.10 | 0.0 | -3.0 | 3.1E-03 | 1.7E-01 | 1.0E+00 |
| MPNI-pz | P43251 | BTD | Biotinidase | -0.08 | 0.0 | -2.1 | 3.2E-02 | 4.4E-01 | 1.0E+00 |
| MPNI-pz | P54289 | CACNA2D1 | Voltage-dependent calcium channel subunit alpha-2/delta-1 | -0.08 | 0.0 | -2.3 | 2.4E-02 | 3.8E-01 | 1.0E+00 |
| MPNI-pz | P98160 | HSPG2 | Basement membrane-specific heparan sulfate proteoglycan core protein | -0.08 | 0.0 | -2.4 | 1.9E-02 | 3.4E-01 | 1.0E+00 |
| MPNI-pz | Q01459 | CTBS | Di-N-acetylchitobiase | 0.07 | 0.0 | 2.0 | 4.7E-02 | 4.7E-01 | 1.0E+00 |
| MPNI-pz | Q07954 | LRP1 | Prolow-density lipoprotein receptor-related protein 1 | -0.08 | 0.0 | -2.2 | 2.5E-02 | 3.8E-01 | 1.0E+00 |
| MPNI-pz | Q12860 | CNTN1 | Contactin-1 | -0.11 | 0.0 | -3.1 | 2.2E-03 | 1.4E-01 | 9.8E-01 |
| MPNI-pz | Q14126 | DSG2 | Desmoglein-2 | -0.10 | 0.0 | -2.8 | 4.6E-03 | 1.8E-01 | 1.0E+00 |
| MPNI-pz | Q14515 | SPARCL1 | SPARC-like protein 1 | -0.10 | 0.0 | -2.9 | 4.0E-03 | 1.8E-01 | 1.0E+00 |
| MPNI-pz | Q15828 | CST6 | Cystatin-M | -0.08 | 0.0 | -2.2 | 2.6E-02 | 3.8E-01 | 1.0E+00 |
| MPNI-pz | Q6YHK3 | CD109 | CD109 antigen | -0.07 | 0.0 | -2.0 | 4.4E-02 | 4.7E-01 | 1.0E+00 |
| MPNI-pz | Q76LX8 | ADAMTS13 | A disintegrin and metalloproteinase with thrombospondin motifs 13 | -0.09 | 0.0 | -2.7 | 7.4E-03 | 2.6E-01 | 1.0E+00 |
| MPNI-pz | Q7Z7M0 | MEGF8 | Multiple epidermal growth factor-like domains protein 8 | -0.09 | 0.0 | -2.5 | 1.3E-02 | 2.9E-01 | 1.0E+00 |
| MPNI-pz | Q99784 | OLFM1 | Noelin | -0.10 | 0.0 | -2.9 | 3.5E-03 | 1.7E-01 | 1.0E+00 |
| MPNI-pz | Q99969 | RARRES2 | Retinoic acid receptor responder protein 2 | 0.11 | 0.0 | 3.5 | 4.4E-04 | 4.8E-02 | 1.9E-01 |
| MPNI-pz | Q99983 | OMD | Osteomodulin | -0.10 | 0.0 | -3.2 | 1.3E-03 | 9.8E-02 | 5.9E-01 |
| MPNI-pz | Q9NPH3 | IL1RAP | Interleukin-1 receptor accessory protein | -0.09 | 0.0 | -2.5 | 1.3E-02 | 2.9E-01 | 1.0E+00 |
| MPNI-pz | Q9Y5Y7 | LYVE1 | Lymphatic vessel endothelial hyaluronic acid receptor 1 | -0.09 | 0.0 | -2.6 | 1.0E-02 | 2.9E-01 | 1.0E+00 |
| MPNI-pz | Q9Y6R7 | FCGBP | IgGFc-binding protein | -0.18 | 0.0 | -4.8 | 1.9E-06 | 8.3E-04 | 8.3E-04 |
| MPNI-p12p | O75636 | FCN3 | Ficolin-3 | 0.67 | 0.3 | 2.2 | 3.0E-02 | 7.3E-01 | 1.0E+00 |
| MPNI-p12p | P00748 | F12 | Coagulation factor XII | -0.69 | 0.3 | -2.2 | 3.0E-02 | 7.3E-01 | 1.0E+00 |
| MPNI-p12p | P01008 | SERPINC1 | Antithrombin-III | 0.63 | 0.3 | 2.1 | 3.8E-02 | 8.3E-01 | 1.0E+00 |
| MPNI-p12p | P02452 | COL1A1 | Collagen alpha-1(I) chain | 0.71 | 0.3 | 2.6 | 1.0E-02 | 7.1E-01 | 1.0E+00 |
| MPNI-p12p | P04278 | SHBG | Sex hormone-binding globulin | -0.93 | 0.4 | -2.4 | 1.5E-02 | 7.1E-01 | 1.0E+00 |
| MPNI-p12p | P05160 | F13B | Coagulation factor XIII B chain | 0.63 | 0.3 | 2.2 | 2.5E-02 | 7.3E-01 | 1.0E+00 |
| MPNI-p12p | P06727 | APOA4 | Apolipoprotein A-IV | 0.59 | 0.3 | 2.0 | 4.7E-02 | 8.3E-01 | 1.0E+00 |

| | | | | | | | | | |
|---|---|---|---|---|---|---|---|---|---|
| MPNI-p12p | P08571 | CD14 | Monocyte differentiation antigen CD14 | 0.92 | 0.3 | 3.4 | 6.1E-04 | 2.7E-01 | 2.7E-01 |
| MPNI-p12p | P09172 | DBH | Dopamine beta-hydroxylase | -0.92 | 0.3 | -3.0 | 3.1E-03 | 6.3E-01 | 1.0E+00 |
| MPNI-p12p | P11047 | LAMC1 | Laminin subunit gamma-1 | -0.61 | 0.3 | -2.3 | 2.3E-02 | 7.3E-01 | 1.0E+00 |
| MPNI-p12p | P12259 | F5 | Coagulation factor V | 0.69 | 0.3 | 2.3 | 1.9E-02 | 7.1E-01 | 1.0E+00 |
| MPNI-p12p | P19021 | PAM | Peptidyl-glycine alpha-amidating monooxygenase | 0.73 | 0.3 | 2.5 | 1.3E-02 | 7.1E-01 | 1.0E+00 |
| MPNI-p12p | P20023 | CR2 | Complement receptor type 2 | 0.81 | 0.3 | 2.8 | 6.0E-03 | 6.5E-01 | 1.0E+00 |
| MPNI-p12p | P20742 | PZP | Pregnancy zone protein | -0.84 | 0.3 | -2.5 | 1.2E-02 | 7.1E-01 | 1.0E+00 |
| MPNI-p12p | P43121 | MCAM | Cell surface glycoprotein MUC18 | 0.56 | 0.3 | 2.0 | 4.3E-02 | 8.3E-01 | 1.0E+00 |
| MPNI-p12p | P43652 | AFM | Afamin | 0.66 | 0.3 | 2.4 | 1.9E-02 | 7.1E-01 | 1.0E+00 |
| MPNI-p12p | P49913 | CAMP | Cathelicidin antimicrobial peptide | -0.61 | 0.3 | -2.2 | 3.0E-02 | 7.3E-01 | 1.0E+00 |
| MPNI-p12p | Q08380 | LGALS3BP | Galectin-3-binding protein | -0.74 | 0.3 | -2.4 | 1.7E-02 | 7.1E-01 | 1.0E+00 |
| MPNI-p12p | Q15848 | ADIPOQ | Adiponectin | -0.57 | 0.3 | -2.1 | 4.0E-02 | 8.3E-01 | 1.0E+00 |
| MPNI-p12p | Q99969 | RARRES2 | Retinoic acid receptor responder protein 2 | 0.54 | 0.3 | 2.1 | 3.5E-02 | 8.0E-01 | 1.0E+00 |
| MPNI-p12p | Q99983 | OMD | Osteomodulin | -0.69 | 0.3 | -2.6 | 9.2E-03 | 7.1E-01 | 1.0E+00 |
| MPNI-p12p | Q9UBX1 | CTSF | Cathepsin F | -0.58 | 0.3 | -2.2 | 2.9E-02 | 7.3E-01 | 1.0E+00 |
| MPNI-p12p | Q9Y6R7 | FCGBP | IgGFc-binding protein | -0.85 | 0.3 | -2.9 | 4.3E-03 | 6.3E-01 | 1.0E+00 |
| MPNI-s14p | O00592 | PODXL | Podocalyxin | -0.85 | 0.3 | -3.1 | 1.9E-03 | 1.2E-01 | 8.2E-01 |
| MPNI-s14p | O14786 | NRP1 | Neuropilin-1 | -0.70 | 0.3 | -2.7 | 6.3E-03 | 1.6E-01 | 1.0E+00 |
| MPNI-s14p | O15204 | ADAMDEC1 | ADAM DEC1 | -0.56 | 0.3 | -2.2 | 3.2E-02 | 2.3E-01 | 1.0E+00 |
| MPNI-s14p | O43866 | CD5L | CD5 antigen-like | -0.56 | 0.3 | -2.1 | 3.4E-02 | 2.4E-01 | 1.0E+00 |
| MPNI-s14p | P00746 | CFD | Complement factor D | -0.63 | 0.3 | -2.2 | 2.7E-02 | 2.2E-01 | 1.0E+00 |
| MPNI-s14p | P02656 | APOC3 | Apolipoprotein C-III | 0.56 | 0.3 | 2.2 | 3.1E-02 | 2.3E-01 | 1.0E+00 |
| MPNI-s14p | P02741 | CRP | C-reactive protein | 0.56 | 0.3 | 2.0 | 4.3E-02 | 2.8E-01 | 1.0E+00 |
| MPNI-s14p | P02743 | APCS | Serum amyloid P-component | 0.64 | 0.3 | 2.2 | 2.6E-02 | 2.2E-01 | 1.0E+00 |
| MPNI-s14p | P02747 | C1QC | Complement C1q subcomponent subunit C | -0.77 | 0.3 | -3.0 | 3.2E-03 | 1.5E-01 | 1.0E+00 |
| MPNI-s14p | P02765 | AHSG | Alpha-2-HS-glycoprotein | -0.57 | 0.3 | -2.2 | 2.8E-02 | 2.2E-01 | 1.0E+00 |
| MPNI-s14p | P05019 | IGF1 | Insulin-like growth factor I | -0.74 | 0.3 | -2.7 | 7.0E-03 | 1.6E-01 | 1.0E+00 |
| MPNI-s14p | P05452 | CLEC3B | Tetranectin | -0.67 | 0.3 | -2.3 | 2.0E-02 | 2.2E-01 | 1.0E+00 |
| MPNI-s14p | P05543 | SERPINA7 | Thyroxine-binding globulin | 0.65 | 0.3 | 2.2 | 2.5E-02 | 2.2E-01 | 1.0E+00 |
| MPNI-s14p | P06396 | GSN | Gelsolin | -0.65 | 0.3 | -2.3 | 1.9E-02 | 2.2E-01 | 1.0E+00 |
| MPNI-s14p | P07911 | UMOD | Uromodulin | -0.54 | 0.3 | -2.1 | 3.9E-02 | 2.6E-01 | 1.0E+00 |
| MPNI-s14p | P08195 | SLC3A2 | 4F2 cell-surface antigen heavy chain | -0.70 | 0.3 | -2.6 | 1.1E-02 | 2.1E-01 | 1.0E+00 |
| MPNI-s14p | P08709 | F7 | Coagulation factor VII | 0.56 | 0.3 | 2.2 | 2.7E-02 | 2.2E-01 | 1.0E+00 |
| MPNI-s14p | P09172 | DBH | Dopamine beta-hydroxylase | -0.62 | 0.3 | -2.2 | 2.6E-02 | 2.2E-01 | 1.0E+00 |
| MPNI-s14p | P0C0L4 | C4A | Complement C4-A | 0.62 | 0.3 | 2.3 | 2.3E-02 | 2.2E-01 | 1.0E+00 |
| MPNI-s14p | P0C0L5 | C4B | Complement C4-B | 1.08 | 0.3 | 4.1 | 4.4E-05 | 1.9E-02 | 1.9E-02 |

| | | | | | | | | | |
|---|---|---|---|---|---|---|---|---|---|
| MPNI-s14p | P10643 | C7 | Complement component C7 | -0.61 | 0.3 | -2.2 | 2.8E-02 | 2.2E-01 | 1.0E+00 |
| MPNI-s14p | P10646 | TFPI | Tissue factor pathway inhibitor | -0.58 | 0.3 | -2.2 | 2.8E-02 | 2.2E-01 | 1.0E+00 |
| MPNI-s14p | P10721 | KIT | Mast/stem cell growth factor receptor Kit | -0.62 | 0.3 | -2.4 | 1.9E-02 | 2.2E-01 | 1.0E+00 |
| MPNI-s14p | P12821 | ACE | Angiotensin-converting enzyme | -0.57 | 0.3 | -2.1 | 3.8E-02 | 2.6E-01 | 1.0E+00 |
| MPNI-s14p | P13598 | ICAM2 | Intercellular adhesion molecule 2 | -0.58 | 0.3 | -2.2 | 2.7E-02 | 2.2E-01 | 1.0E+00 |
| MPNI-s14p | P14151 | SELL | L-selectin | -0.52 | 0.3 | -2.0 | 4.5E-02 | 2.8E-01 | 1.0E+00 |
| MPNI-s14p | P14618 | PKM | Pyruvate kinase PKM | -0.65 | 0.3 | -2.5 | 1.4E-02 | 2.1E-01 | 1.0E+00 |
| MPNI-s14p | P15144 | ANPEP | Aminopeptidase N | -0.62 | 0.3 | -2.4 | 1.7E-02 | 2.2E-01 | 1.0E+00 |
| MPNI-s14p | P16035 | TIMP2 | Metalloproteinase inhibitor 2 | -0.52 | 0.3 | -2.0 | 4.4E-02 | 2.8E-01 | 1.0E+00 |
| MPNI-s14p | P17936 | IGFBP3 | Insulin-like growth factor-binding protein 3 | -0.78 | 0.3 | -2.9 | 4.1E-03 | 1.5E-01 | 1.0E+00 |
| MPNI-s14p | P19823 | ITIH2 | Inter-alpha-trypsin inhibitor heavy chain H2 | -0.60 | 0.3 | -2.3 | 2.3E-02 | 2.2E-01 | 1.0E+00 |
| MPNI-s14p | P19827 | ITIH1 | Inter-alpha-trypsin inhibitor heavy chain H1 | -0.63 | 0.3 | -2.4 | 1.6E-02 | 2.2E-01 | 1.0E+00 |
| MPNI-s14p | P20023 | CR2 | Complement receptor type 2 | -0.62 | 0.3 | -2.3 | 2.4E-02 | 2.2E-01 | 1.0E+00 |
| MPNI-s14p | P20742 | PZP | Pregnancy zone protein | 0.69 | 0.3 | 2.2 | 2.8E-02 | 2.2E-01 | 1.0E+00 |
| MPNI-s14p | P23142 | FBLN1 | Fibulin-1 | -0.63 | 0.3 | -2.4 | 1.5E-02 | 2.1E-01 | 1.0E+00 |
| MPNI-s14p | P24593 | IGFBP5 | Insulin-like growth factor-binding protein 5 | -0.72 | 0.3 | -2.8 | 5.8E-03 | 1.6E-01 | 1.0E+00 |
| MPNI-s14p | P28799 | GRN | Progranulin | -0.68 | 0.3 | -2.6 | 9.1E-03 | 1.9E-01 | 1.0E+00 |
| MPNI-s14p | P40189 | IL6ST | Interleukin-6 receptor subunit beta | -0.93 | 0.3 | -3.7 | 2.2E-04 | 3.3E-02 | 9.8E-02 |
| MPNI-s14p | P43121 | MCAM | Cell surface glycoprotein MUC18 | -0.70 | 0.3 | -2.7 | 7.5E-03 | 1.6E-01 | 1.0E+00 |
| MPNI-s14p | P43251 | BTD | Biotinidase | -0.52 | 0.3 | -2.0 | 4.8E-02 | 3.0E-01 | 1.0E+00 |
| MPNI-s14p | P49747 | COMP | Cartilage oligomeric matrix protein | -0.58 | 0.3 | -2.1 | 4.1E-02 | 2.7E-01 | 1.0E+00 |
| MPNI-s14p | P54289 | CACNA2D1 | Voltage-dependent calcium channel subunit alpha-2/delta-1 | -0.67 | 0.3 | -2.6 | 9.7E-03 | 1.9E-01 | 1.0E+00 |
| MPNI-s14p | P55056 | APOC4 | Apolipoprotein C-IV | 0.56 | 0.3 | 2.2 | 3.0E-02 | 2.2E-01 | 1.0E+00 |
| MPNI-s14p | P55290 | CDH13 | Cadherin-13 | -0.87 | 0.3 | -3.3 | 1.1E-03 | 9.3E-02 | 4.6E-01 |
| MPNI-s14p | P62913 | RPL11 | 60S ribosomal protein L11 | -0.67 | 0.3 | -2.5 | 1.1E-02 | 2.1E-01 | 1.0E+00 |
| MPNI-s14p | P98160 | HSPG2 | Basement membrane-specific heparan sulfate proteoglycan core protein | -0.59 | 0.3 | -2.3 | 2.2E-02 | 2.2E-01 | 1.0E+00 |
| MPNI-s14p | Q01469 | FABP5 | Fatty acid-binding protein 5 | -0.53 | 0.3 | -2.1 | 3.5E-02 | 2.4E-01 | 1.0E+00 |
| MPNI-s14p | Q04756 | HGFAC | Hepatocyte growth factor activator | -0.60 | 0.3 | -2.2 | 3.0E-02 | 2.2E-01 | 1.0E+00 |
| MPNI-s14p | Q07954 | LRP1 | Prolow-density lipoprotein receptor-related protein 1 | -0.65 | 0.3 | -2.5 | 1.4E-02 | 2.1E-01 | 1.0E+00 |
| MPNI-s14p | Q12805 | EFEMP1 | EGF-containing fibulin-like extracellular matrix protein 1 | -0.58 | 0.3 | -2.2 | 2.7E-02 | 2.2E-01 | 1.0E+00 |

| | | | | | | | | | |
|---|---|---|---|---|---|---|---|---|---|
| MPNI-s14p | Q12860 | CNTN1 | Contactin-1 | -0.65 | 0.3 | -2.5 | 1.3E-02 | 2.1E-01 | 1.0E+00 |
| MPNI-s14p | Q13449 | LSAMP | Limbic system-associated membrane protein | -0.76 | 0.3 | -2.9 | 4.2E-03 | 1.5E-01 | 1.0E+00 |
| MPNI-s14p | Q14126 | DSG2 | Desmoglein-2 | -0.69 | 0.3 | -2.8 | 6.0E-03 | 1.6E-01 | 1.0E+00 |
| MPNI-s14p | Q14515 | SPARCL1 | SPARC-like protein 1 | -0.99 | 0.3 | -3.9 | 1.1E-04 | 2.4E-02 | 4.9E-02 |
| MPNI-s14p | Q14956 | GPNMB | Transmembrane glycoprotein NMB | -0.81 | 0.3 | -3.1 | 1.9E-03 | 1.2E-01 | 8.4E-01 |
| MPNI-s14p | Q6EMK4 | VASN | Vasorin | -0.64 | 0.3 | -2.3 | 2.0E-02 | 2.2E-01 | 1.0E+00 |
| MPNI-s14p | Q6UX71 | PLXDC2 | Plexin domain-containing protein 2 | -0.61 | 0.3 | -2.2 | 2.6E-02 | 2.2E-01 | 1.0E+00 |
| MPNI-s14p | Q6YHK3 | CD109 | CD109 antigen | -0.63 | 0.3 | -2.4 | 1.8E-02 | 2.2E-01 | 1.0E+00 |
| MPNI-s14p | Q7Z7M0 | MEGF8 | Multiple epidermal growth factor-like domains protein 8 | -0.68 | 0.3 | -2.5 | 1.3E-02 | 2.1E-01 | 1.0E+00 |
| MPNI-s14p | Q86TH1 | ADAMTSL2 | ADAMTS-like protein 2 | -0.79 | 0.3 | -3.1 | 2.3E-03 | 1.3E-01 | 1.0E+00 |
| MPNI-s14p | Q86U17 | SERPINA11 | Serpin A11 | -0.62 | 0.3 | -2.4 | 1.8E-02 | 2.2E-01 | 1.0E+00 |
| MPNI-s14p | Q8IUL8 | CILP2 | Cartilage intermediate layer protein 2 | -0.70 | 0.3 | -2.8 | 5.9E-03 | 1.6E-01 | 1.0E+00 |
| MPNI-s14p | Q99784 | OLFM1 | Noelin | -0.75 | 0.3 | -2.9 | 3.7E-03 | 1.5E-01 | 1.0E+00 |
| MPNI-s14p | Q99983 | OMD | Osteomodulin | -0.61 | 0.2 | -2.4 | 1.5E-02 | 2.1E-01 | 1.0E+00 |
| MPNI-s14p | Q9H4G4 | GLIPR2 | Golgi-associated plant pathogenesis-related protein 1 | -0.55 | 0.3 | -2.2 | 3.0E-02 | 2.2E-01 | 1.0E+00 |
| MPNI-s14p | Q9NPH3 | IL1RAP | Interleukin-1 receptor accessory protein | -0.92 | 0.3 | -3.4 | 8.2E-04 | 9.1E-02 | 3.6E-01 |
| MPNI-s14p | Q9NPR2 | SEMA4B | Semaphorin-4B | -0.73 | 0.3 | -2.7 | 7.3E-03 | 1.6E-01 | 1.0E+00 |
| MPNI-s14p | Q9NY15 | STAB1 | Stabilin-1 | -0.62 | 0.3 | -2.5 | 1.3E-02 | 2.1E-01 | 1.0E+00 |
| MPNI-s14p | Q9Y251 | HPSE | Heparanase | -0.53 | 0.3 | -2.1 | 3.8E-02 | 2.6E-01 | 1.0E+00 |
| MPNI-s14p | Q9Y5Y7 | LYVE1 | Lymphatic vessel endothelial hyaluronic acid receptor 1 | -0.62 | 0.3 | -2.4 | 1.7E-02 | 2.2E-01 | 1.0E+00 |
| MPNI-s14p | Q9Y6R7 | FCGBP | IgGFc-binding protein | -0.75 | 0.3 | -2.7 | 7.3E-03 | 1.6E-01 | 1.0E+00 |
| MPNI-s17p | O15204 | ADAMDEC1 | ADAM DEC1 | -0.75 | 0.3 | -2.8 | 5.1E-03 | 3.1E-01 | 1.0E+00 |
| MPNI-s17p | O95497 | VNN1 | Pantetheinase | 0.57 | 0.3 | 2.2 | 2.8E-02 | 5.2E-01 | 1.0E+00 |
| MPNI-s17p | P02743 | APCS | Serum amyloid P-component | 0.66 | 0.3 | 2.3 | 2.1E-02 | 4.9E-01 | 1.0E+00 |
| MPNI-s17p | P03950 | ANG | Angiogenin | 0.68 | 0.3 | 2.5 | 1.2E-02 | 3.5E-01 | 1.0E+00 |
| MPNI-s17p | P05062 | ALDOB | Fructose-bisphosphate aldolase B | 0.58 | 0.3 | 2.2 | 2.9E-02 | 5.2E-01 | 1.0E+00 |
| MPNI-s17p | P05556 | ITGB1 | Integrin beta-1 | -0.54 | 0.3 | -2.1 | 3.8E-02 | 5.2E-01 | 1.0E+00 |
| MPNI-s17p | P07339 | CTSD | Cathepsin D | 0.56 | 0.3 | 2.1 | 3.4E-02 | 5.2E-01 | 1.0E+00 |
| MPNI-s17p | P07911 | UMOD | Uromodulin | -0.70 | 0.3 | -2.7 | 7.1E-03 | 3.1E-01 | 1.0E+00 |
| MPNI-s17p | P07942 | LAMB1 | Laminin subunit beta-1 | -0.53 | 0.3 | -2.1 | 3.8E-02 | 5.2E-01 | 1.0E+00 |
| MPNI-s17p | P09172 | DBH | Dopamine beta-hydroxylase | -0.54 | 0.3 | -2.0 | 4.8E-02 | 5.7E-01 | 1.0E+00 |
| MPNI-s17p | P10586 | PTPRF | Receptor-type tyrosine-protein phosphatase F | -0.49 | 0.2 | -2.0 | 4.5E-02 | 5.7E-01 | 1.0E+00 |

| | | | | | | | | | |
|---|---|---|---|---|---|---|---|---|---|
| MPNI-s17p | P10721 | KIT | Mast/stem cell growth factor receptor Kit | -0.97 | 0.3 | -3.7 | 2.8E-04 | 6.2E-02 | 1.2E-01 |
| MPNI-s17p | P11279 | LAMP1 | Lysosome-associated membrane glycoprotein 1 | 0.54 | 0.3 | 2.1 | 3.9E-02 | 5.2E-01 | 1.0E+00 |
| MPNI-s17p | P11362 | FGFR1 | Fibroblast growth factor receptor 1 | -0.52 | 0.3 | -2.0 | 4.8E-02 | 5.7E-01 | 1.0E+00 |
| MPNI-s17p | P12821 | ACE | Angiotensin-converting enzyme | -0.74 | 0.3 | -2.7 | 6.6E-03 | 3.1E-01 | 1.0E+00 |
| MPNI-s17p | P12955 | PEPD | Xaa-Pro dipeptidase | -0.57 | 0.3 | -2.2 | 2.9E-02 | 5.2E-01 | 1.0E+00 |
| MPNI-s17p | P15151 | PVR | Poliovirus receptor | 0.59 | 0.3 | 2.3 | 2.2E-02 | 4.9E-01 | 1.0E+00 |
| MPNI-s17p | P16035 | TIMP2 | Metalloproteinase inhibitor 2 | -0.63 | 0.3 | -2.4 | 1.6E-02 | 4.0E-01 | 1.0E+00 |
| MPNI-s17p | P23142 | FBLN1 | Fibulin-1 | -1.04 | 0.3 | -4.1 | 5.0E-05 | 2.2E-02 | 2.2E-02 |
| MPNI-s17p | P35916 | FLT4 | Vascular endothelial growth factor receptor 3 | 0.66 | 0.3 | 2.5 | 1.3E-02 | 3.5E-01 | 1.0E+00 |
| MPNI-s17p | P98160 | HSPG2 | Basement membrane-specific heparan sulfate proteoglycan core protein | -0.54 | 0.3 | -2.1 | 4.0E-02 | 5.2E-01 | 1.0E+00 |
| MPNI-s17p | Q00610 | CLTC | Clathrin heavy chain 1 | -0.53 | 0.2 | -2.2 | 3.1E-02 | 5.2E-01 | 1.0E+00 |
| MPNI-s17p | Q01469 | FABP5 | Fatty acid-binding protein 5 | -0.55 | 0.3 | -2.1 | 3.6E-02 | 5.2E-01 | 1.0E+00 |
| MPNI-s17p | Q04695 | KRT17 | Keratin, type I cytoskeletal 17 | 0.57 | 0.3 | 2.3 | 2.5E-02 | 5.2E-01 | 1.0E+00 |
| MPNI-s17p | Q12805 | EFEMP1 | EGF-containing fibulin-like extracellular matrix protein 1 | -0.80 | 0.3 | -3.1 | 2.0E-03 | 1.7E-01 | 8.6E-01 |
| MPNI-s17p | Q12860 | CNTN1 | Contactin-1 | -0.85 | 0.3 | -3.2 | 1.6E-03 | 1.7E-01 | 7.1E-01 |
| MPNI-s17p | Q14515 | SPARCL1 | SPARC-like protein 1 | -0.64 | 0.3 | -2.5 | 1.3E-02 | 3.5E-01 | 1.0E+00 |
| MPNI-s17p | Q15063 | POSTN | Periostin | -0.69 | 0.3 | -2.7 | 7.1E-03 | 3.1E-01 | 1.0E+00 |
| MPNI-s17p | Q6YHK3 | CD109 | CD109 antigen | -0.70 | 0.3 | -2.6 | 9.0E-03 | 3.5E-01 | 1.0E+00 |
| MPNI-s17p | Q86TH1 | ADAMTSL2 | ADAMTS-like protein 2 | -0.56 | 0.3 | -2.1 | 3.6E-02 | 5.2E-01 | 1.0E+00 |
| MPNI-s17p | Q86UD1 | OAF | Out at first protein homolog | 0.63 | 0.3 | 2.5 | 1.3E-02 | 3.5E-01 | 1.0E+00 |
| MPNI-s17p | Q92820 | GGH | Gamma-glutamyl hydrolase | 0.65 | 0.3 | 2.5 | 1.3E-02 | 3.5E-01 | 1.0E+00 |
| MPNI-s17p | Q99969 | RARRES2 | Retinoic acid receptor responder protein 2 | 0.53 | 0.3 | 2.1 | 3.9E-02 | 5.2E-01 | 1.0E+00 |
| MPNI-s17p | Q9NPH3 | IL1RAP | Interleukin-1 receptor accessory protein | -0.92 | 0.3 | -3.3 | 1.1E-03 | 1.6E-01 | 4.7E-01 |
| MPNI-s17p | Q9UJJ9 | GNPTG | N-acetylglucosamine-1-phosphotransferase subunit gamma | -0.53 | 0.2 | -2.2 | 3.1E-02 | 5.2E-01 | 1.0E+00 |
| MPNI-s17p | Q9Y5Y7 | LYVE1 | Lymphatic vessel endothelial hyaluronic acid receptor 1 | -0.66 | 0.3 | -2.5 | 1.2E-02 | 3.5E-01 | 1.0E+00 |
| MPNI-s17p | Q9Y6R7 | FCGBP | IgGFc-binding protein | -0.79 | 0.3 | -2.8 | 5.3E-03 | 3.1E-01 | 1.0E+00 |
| MPNI-t12p | O14791 | APOL1 | Apolipoprotein L1 | 0.91 | 0.4 | 2.1 | 3.2E-02 | 6.2E-01 | 1.0E+00 |
| MPNI-t12p | O75636 | FCN3 | Ficolin-3 | 0.91 | 0.4 | 2.2 | 3.0E-02 | 6.1E-01 | 1.0E+00 |
| MPNI-t12p | O95445 | APOM | Apolipoprotein M | 0.95 | 0.4 | 2.5 | 1.3E-02 | 4.8E-01 | 1.0E+00 |
| MPNI-t12p | P00734 | F2 | Prothrombin | 1.00 | 0.4 | 2.7 | 8.1E-03 | 4.5E-01 | 1.0E+00 |

| | | | | | | | | | |
|---|---|---|---|---|---|---|---|---|---|
| MPNI-t12p | P00742 | F10 | Coagulation factor X | 0.83 | 0.4 | 2.1 | 3.9E-02 | 6.2E-01 | 1.0E+00 |
| MPNI-t12p | P02652 | APOA2 | Apolipoprotein A-II | 1.01 | 0.4 | 2.6 | 9.0E-03 | 4.5E-01 | 1.0E+00 |
| MPNI-t12p | P02749 | APOH | Beta-2-glycoprotein 1 | 0.83 | 0.4 | 2.0 | 4.8E-02 | 6.2E-01 | 1.0E+00 |
| MPNI-t12p | P04070 | PROC | Vitamin K-dependent protein C | 0.85 | 0.4 | 2.2 | 2.5E-02 | 5.6E-01 | 1.0E+00 |
| MPNI-t12p | P05067 | APP | Amyloid-beta precursor protein | 0.95 | 0.4 | 2.6 | 8.7E-03 | 4.5E-01 | 1.0E+00 |
| MPNI-t12p | P05160 | F13B | Coagulation factor XIII B chain | 0.79 | 0.4 | 2.1 | 3.4E-02 | 6.2E-01 | 1.0E+00 |
| MPNI-t12p | P05546 | SERPIND1 | Heparin cofactor 2 | 1.04 | 0.4 | 2.7 | 6.4E-03 | 4.5E-01 | 1.0E+00 |
| MPNI-t12p | P07911 | UMOD | Uromodulin | -0.95 | 0.4 | -2.5 | 1.1E-02 | 4.8E-01 | 1.0E+00 |
| MPNI-t12p | P07996 | THBS1 | Thrombospondin-1 | 0.76 | 0.4 | 2.0 | 4.9E-02 | 6.2E-01 | 1.0E+00 |
| MPNI-t12p | P08571 | CD14 | Monocyte differentiation antigen CD14 | 0.96 | 0.4 | 2.6 | 9.2E-03 | 4.5E-01 | 1.0E+00 |
| MPNI-t12p | P09172 | DBH | Dopamine beta-hydroxylase | -1.02 | 0.4 | -2.4 | 1.6E-02 | 5.1E-01 | 1.0E+00 |
| MPNI-t12p | P09486 | SPARC | SPARC | 1.06 | 0.4 | 2.6 | 8.7E-03 | 4.5E-01 | 1.0E+00 |
| MPNI-t12p | P10720 | PF4V1 | Platelet factor 4 variant | 0.77 | 0.4 | 2.1 | 3.8E-02 | 6.2E-01 | 1.0E+00 |
| MPNI-t12p | P14618 | PKM | Pyruvate kinase PKM | -0.88 | 0.4 | -2.3 | 2.0E-02 | 5.2E-01 | 1.0E+00 |
| MPNI-t12p | P19021 | PAM | Peptidyl-glycine alpha-amidating monooxygenase | 0.89 | 0.4 | 2.3 | 2.1E-02 | 5.2E-01 | 1.0E+00 |
| MPNI-t12p | P23142 | FBLN1 | Fibulin-1 | -0.73 | 0.4 | -2.0 | 4.9E-02 | 6.2E-01 | 1.0E+00 |
| MPNI-t12p | P26927 | MST1 | Hepatocyte growth factor-like protein | 0.88 | 0.4 | 2.4 | 1.8E-02 | 5.1E-01 | 1.0E+00 |
| MPNI-t12p | P27169 | PON1 | Serum paraoxonase/arylesterase 1 | 0.91 | 0.4 | 2.3 | 2.1E-02 | 5.2E-01 | 1.0E+00 |
| MPNI-t12p | P43652 | AFM | Afamin | 1.20 | 0.4 | 3.1 | 2.1E-03 | 4.5E-01 | 9.0E-01 |
| MPNI-t12p | P48740 | MASP1 | Mannan-binding lectin serine protease 1 | 0.96 | 0.4 | 2.5 | 1.2E-02 | 4.8E-01 | 1.0E+00 |
| MPNI-t12p | P68871 | HBB | Hemoglobin subunit beta | -0.79 | 0.4 | -2.2 | 2.6E-02 | 5.6E-01 | 1.0E+00 |
| MPNI-t12p | Q01459 | CTBS | Di-N-acetylchitobiase | 1.33 | 0.4 | 3.6 | 3.9E-04 | 1.7E-01 | 1.7E-01 |
| MPNI-t12p | Q14766 | LTBP1 | Latent-transforming growth factor beta-binding protein 1 | 0.91 | 0.4 | 2.4 | 1.8E-02 | 5.1E-01 | 1.0E+00 |
| MPNI-t12p | Q86SQ4 | ADGRG6 | Adhesion G-protein coupled receptor G6 | -0.81 | 0.4 | -2.0 | 4.2E-02 | 6.2E-01 | 1.0E+00 |
| MPNI-t12p | Q96EE4 | CCDC126 | Coiled-coil domain-containing protein 126 | 0.94 | 0.4 | 2.7 | 8.1E-03 | 4.5E-01 | 1.0E+00 |
| MPNI-t12p | Q96NZ9 | PRAP1 | Proline-rich acidic protein 1 | 0.74 | 0.4 | 2.1 | 4.1E-02 | 6.2E-01 | 1.0E+00 |
| MPNI-t12p | Q99983 | OMD | Osteomodulin | -0.77 | 0.4 | -2.2 | 3.0E-02 | 6.1E-01 | 1.0E+00 |
| MPNI-t14p | O95497 | VNN1 | Pantetheinase | 0.82 | 0.4 | 2.1 | 4.0E-02 | 1.0E+00 | 1.0E+00 |
| MPNI-t14p | P02654 | APOC1 | Apolipoprotein C-I | -0.82 | 0.4 | -2.3 | 2.5E-02 | 1.0E+00 | 1.0E+00 |
| MPNI-t14p | P05019 | IGF1 | Insulin-like growth factor I | -0.97 | 0.4 | -2.6 | 1.1E-02 | 1.0E+00 | 1.0E+00 |
| MPNI-t14p | P05164 | MPO | Myeloperoxidase | 0.82 | 0.4 | 2.1 | 3.5E-02 | 1.0E+00 | 1.0E+00 |
| MPNI-t14p | P08571 | CD14 | Monocyte differentiation antigen CD14 | 0.99 | 0.4 | 2.7 | 7.0E-03 | 1.0E+00 | 1.0E+00 |
| MPNI-t14p | P11597 | CETP | Cholesteryl ester transfer protein | -0.74 | 0.4 | -2.0 | 4.8E-02 | 1.0E+00 | 1.0E+00 |
| MPNI-t14p | P15169 | CPN1 | Carboxypeptidase N catalytic chain | -0.88 | 0.4 | -2.0 | 5.0E-02 | 1.0E+00 | 1.0E+00 |

| | | | | | | | | | |
|---|---|---|---|---|---|---|---|---|---|
| MPNI-t14p | Q01459 | CTBS | Di-N-acetylchitobiase | 1.29 | 0.4 | 3.3 | 9.9E-04 | 4.3E-01 | 4.3E-01 |
| MPNI-t14p | Q13103 | SPP2 | Secreted phosphoprotein 24 | -1.03 | 0.4 | -2.4 | 1.8E-02 | 1.0E+00 | 1.0E+00 |
| MPNI-t14p | Q96S96 | PEBP4 | Phosphatidylethanolamine-binding protein 4 | -0.90 | 0.4 | -2.2 | 3.2E-02 | 1.0E+00 | 1.0E+00 |
| MPNI-t14p | Q99983 | OMD | Osteomodulin | -0.94 | 0.4 | -2.5 | 1.3E-02 | 1.0E+00 | 1.0E+00 |
| MPNI-t14p | Q9HDC9 | APMAP | Adipocyte plasma membrane-associated protein | -0.86 | 0.4 | -2.2 | 2.8E-02 | 1.0E+00 | 1.0E+00 |
| MPNI-t14p | Q9UNN8 | PROCR | Endothelial protein C receptor | -0.95 | 0.4 | -2.3 | 1.9E-02 | 1.0E+00 | 1.0E+00 |
| MPNI-co14p | O75144 | ICOSLG | ICOS ligand | -0.71 | 0.3 | -2.3 | 2.3E-02 | 6.6E-01 | 1.0E+00 |
| MPNI-co14p | O75882 | ATRN | Attractin | -0.66 | 0.3 | -2.1 | 4.0E-02 | 6.6E-01 | 1.0E+00 |
| MPNI-co14p | O95479 | H6PD | GDH/6PGL endoplasmic bifunctional protein | -0.71 | 0.3 | -2.2 | 2.8E-02 | 6.6E-01 | 1.0E+00 |
| MPNI-co14p | P01031 | C5 | Complement C5 | 0.70 | 0.3 | 2.2 | 2.9E-02 | 6.6E-01 | 1.0E+00 |
| MPNI-co14p | P01344 | IGF2 | Insulin-like growth factor II | -0.72 | 0.3 | -2.3 | 2.4E-02 | 6.6E-01 | 1.0E+00 |
| MPNI-co14p | P05121 | SERPINE1 | Plasminogen activator inhibitor 1 | -0.76 | 0.3 | -2.4 | 1.5E-02 | 6.6E-01 | 1.0E+00 |
| MPNI-co14p | P05362 | ICAM1 | Intercellular adhesion molecule 1 | 0.64 | 0.3 | 2.0 | 4.5E-02 | 6.6E-01 | 1.0E+00 |
| MPNI-co14p | P06727 | APOA4 | Apolipoprotein A-IV | 0.68 | 0.3 | 2.0 | 4.4E-02 | 6.6E-01 | 1.0E+00 |
| MPNI-co14p | P07911 | UMOD | Uromodulin | -0.70 | 0.3 | -2.2 | 2.9E-02 | 6.6E-01 | 1.0E+00 |
| MPNI-co14p | P08571 | CD14 | Monocyte differentiation antigen CD14 | 1.02 | 0.3 | 3.2 | 1.4E-03 | 2.1E-01 | 6.3E-01 |
| MPNI-co14p | P12111 | COL6A3 | Collagen alpha-3(VI) chain | -0.62 | 0.3 | -2.0 | 4.3E-02 | 6.6E-01 | 1.0E+00 |
| MPNI-co14p | P19021 | PAM | Peptidyl-glycine alpha-amidating monooxygenase | 0.80 | 0.3 | 2.3 | 1.9E-02 | 6.6E-01 | 1.0E+00 |
| MPNI-co14p | P23142 | FBLN1 | Fibulin-1 | -0.69 | 0.3 | -2.2 | 2.9E-02 | 6.6E-01 | 1.0E+00 |
| MPNI-co14p | P27105 | STOM | Stomatin | 0.67 | 0.3 | 2.1 | 3.7E-02 | 6.6E-01 | 1.0E+00 |
| MPNI-co14p | P27797 | CALR | Calreticulin | 0.63 | 0.3 | 2.0 | 4.7E-02 | 6.6E-01 | 1.0E+00 |
| MPNI-co14p | P35908 | KRT2 | Keratin, type II cytoskeletal 2 epidermal | -0.61 | 0.3 | -2.0 | 4.8E-02 | 6.6E-01 | 1.0E+00 |
| MPNI-co14p | P41222 | PTGDS | Prostaglandin-H2 D-isomerase | -0.76 | 0.3 | -2.4 | 1.9E-02 | 6.6E-01 | 1.0E+00 |
| MPNI-co14p | P48740 | MASP1 | Mannan-binding lectin serine protease 1 | 0.72 | 0.3 | 2.2 | 2.6E-02 | 6.6E-01 | 1.0E+00 |
| MPNI-co14p | P55290 | CDH13 | Cadherin-13 | -0.90 | 0.3 | -2.8 | 5.4E-03 | 4.0E-01 | 1.0E+00 |
| MPNI-co14p | P59666 | DEFA3 | Neutrophil defensin 3 | 0.65 | 0.3 | 2.2 | 3.2E-02 | 6.6E-01 | 1.0E+00 |
| MPNI-co14p | Q07954 | LRP1 | Prolow-density lipoprotein receptor-related protein 1 | -0.64 | 0.3 | -2.0 | 4.7E-02 | 6.6E-01 | 1.0E+00 |
| MPNI-co14p | Q12860 | CNTN1 | Contactin-1 | -1.04 | 0.3 | -3.2 | 1.4E-03 | 2.1E-01 | 6.3E-01 |
| MPNI-co14p | Q14520 | HABP2 | Hyaluronan-binding protein 2 | 0.70 | 0.3 | 2.1 | 3.7E-02 | 6.6E-01 | 1.0E+00 |
| MPNI-co14p | Q6YHK3 | CD109 | CD109 antigen | -0.91 | 0.3 | -2.8 | 4.9E-03 | 4.0E-01 | 1.0E+00 |
| MPNI-co14p | Q76LX8 | ADAMTS13 | A disintegrin and metalloproteinase with thrombospondin motifs 13 | -0.77 | 0.3 | -2.4 | 1.7E-02 | 6.6E-01 | 1.0E+00 |

| | | | | | | | | | |
|---|---|---|---|---|---|---|---|---|---|
| MPNI-co14p | Q96NZ9 | PRAP1 | Proline-rich acidic protein 1 | -0.72 | 0.3 | -2.2 | 2.7E-02 | 6.6E-01 | 1.0E+00 |
| MPNI-co14p | Q99784 | OLFM1 | Noelin | -0.75 | 0.3 | -2.4 | 1.6E-02 | 6.6E-01 | 1.0E+00 |
| MPNI-co14p | Q99983 | OMD | Osteomodulin | -0.63 | 0.3 | -2.1 | 3.9E-02 | 6.6E-01 | 1.0E+00 |
| MPNI-co14p | Q9NTU7 | CBLN4 | Cerebellin-4 | -0.90 | 0.3 | -2.9 | 4.5E-03 | 4.0E-01 | 1.0E+00 |
| MPNI-co14p | Q9Y6R7 | FCGBP | IgGFc-binding protein | -1.34 | 0.3 | -4.0 | 5.9E-05 | 2.6E-02 | 2.6E-02 |
| MPNI-co17p | O00592 | PODXL | Podocalyxin | -0.73 | 0.3 | -2.2 | 3.1E-02 | 3.6E-01 | 1.0E+00 |
| MPNI-co17p | O14786 | NRP1 | Neuropilin-1 | -0.80 | 0.3 | -2.5 | 1.2E-02 | 2.9E-01 | 1.0E+00 |
| MPNI-co17p | O43866 | CD5L | CD5 antigen-like | -1.03 | 0.3 | -3.3 | 1.2E-03 | 1.7E-01 | 5.3E-01 |
| MPNI-co17p | O75882 | ATRN | Attractin | -0.78 | 0.3 | -2.5 | 1.4E-02 | 2.9E-01 | 1.0E+00 |
| MPNI-co17p | O95497 | VNN1 | Pantetheinase | 0.79 | 0.3 | 2.5 | 1.2E-02 | 2.9E-01 | 1.0E+00 |
| MPNI-co17p | P02144 | MB | Myoglobin | -0.69 | 0.3 | -2.1 | 3.3E-02 | 3.6E-01 | 1.0E+00 |
| MPNI-co17p | P02533 | KRT14 | Keratin, type I cytoskeletal 14 | -0.63 | 0.3 | -2.0 | 4.5E-02 | 4.0E-01 | 1.0E+00 |
| MPNI-co17p | P02747 | C1QC | Complement C1q subcomponent subunit C | -1.07 | 0.3 | -3.4 | 6.4E-04 | 1.7E-01 | 2.8E-01 |
| MPNI-co17p | P02753 | RBP4 | Retinol-binding protein 4 | 0.71 | 0.3 | 2.2 | 2.7E-02 | 3.6E-01 | 1.0E+00 |
| MPNI-co17p | P03950 | ANG | Angiogenin | 0.75 | 0.3 | 2.3 | 2.3E-02 | 3.6E-01 | 1.0E+00 |
| MPNI-co17p | P04003 | C4BPA | C4b-binding protein alpha chain | -0.66 | 0.3 | -2.0 | 4.9E-02 | 4.1E-01 | 1.0E+00 |
| MPNI-co17p | P04264 | KRT1 | Keratin, type II cytoskeletal 1 | -0.62 | 0.3 | -2.0 | 4.3E-02 | 4.0E-01 | 1.0E+00 |
| MPNI-co17p | P04746 | AMY2A | Pancreatic alpha-amylase | -0.99 | 0.3 | -3.2 | 1.4E-03 | 1.7E-01 | 5.9E-01 |
| MPNI-co17p | P05546 | SERPIND1 | Heparin cofactor 2 | 0.64 | 0.3 | 2.0 | 4.7E-02 | 4.0E-01 | 1.0E+00 |
| MPNI-co17p | P07333 | CSF1R | Macrophage colony-stimulating factor 1 receptor | -0.80 | 0.3 | -2.5 | 1.1E-02 | 2.9E-01 | 1.0E+00 |
| MPNI-co17p | P07911 | UMOD | Uromodulin | -0.86 | 0.3 | -2.7 | 7.6E-03 | 2.6E-01 | 1.0E+00 |
| MPNI-co17p | P08238 | HSP90AB1 | Heat shock protein HSP 90-beta | -0.87 | 0.3 | -2.8 | 6.0E-03 | 2.4E-01 | 1.0E+00 |
| MPNI-co17p | P09172 | DBH | Dopamine beta-hydroxylase | -0.73 | 0.3 | -2.2 | 2.9E-02 | 3.6E-01 | 1.0E+00 |
| MPNI-co17p | P09960 | LTA4H | Leukotriene A-4 hydrolase | 0.90 | 0.3 | 3.0 | 3.1E-03 | 2.1E-01 | 1.0E+00 |
| MPNI-co17p | P13646 | KRT13 | Keratin, type I cytoskeletal 13 | 0.61 | 0.3 | 2.0 | 4.4E-02 | 4.0E-01 | 1.0E+00 |
| MPNI-co17p | P14314 | PRKCSH | Glucosidase 2 subunit beta | -0.77 | 0.3 | -2.3 | 2.1E-02 | 3.6E-01 | 1.0E+00 |
| MPNI-co17p | P14543 | NID1 | Nidogen-1 | -0.65 | 0.3 | -2.1 | 3.2E-02 | 3.6E-01 | 1.0E+00 |
| MPNI-co17p | P16035 | TIMP2 | Metalloproteinase inhibitor 2 | -0.78 | 0.3 | -2.4 | 1.5E-02 | 2.9E-01 | 1.0E+00 |
| MPNI-co17p | P18206 | VCL | Vinculin | -0.77 | 0.3 | -2.5 | 1.4E-02 | 2.9E-01 | 1.0E+00 |
| MPNI-co17p | P19021 | PAM | Peptidyl-glycine alpha-amidating monooxygenase | 0.74 | 0.3 | 2.2 | 3.0E-02 | 3.6E-01 | 1.0E+00 |
| MPNI-co17p | P19320 | VCAM1 | Vascular cell adhesion protein 1 | -0.63 | 0.3 | -2.0 | 4.6E-02 | 4.0E-01 | 1.0E+00 |
| MPNI-co17p | P19827 | ITIH1 | Inter-alpha-trypsin inhibitor heavy chain H1 | -0.69 | 0.3 | -2.1 | 3.4E-02 | 3.6E-01 | 1.0E+00 |
| MPNI-co17p | P20851 | C4BPB | C4b-binding protein beta chain | -0.76 | 0.3 | -2.2 | 2.6E-02 | 3.6E-01 | 1.0E+00 |
| MPNI-co17p | P23142 | FBLN1 | Fibulin-1 | -0.87 | 0.3 | -2.8 | 5.4E-03 | 2.4E-01 | 1.0E+00 |
| MPNI-co17p | P27487 | DPP4 | Dipeptidyl peptidase 4 | -0.72 | 0.3 | -2.2 | 3.1E-02 | 3.6E-01 | 1.0E+00 |

| | | | | | | | | | |
|---|---|---|---|---|---|---|---|---|---|
| MPNI-co17p | P41222 | PTGDS | Prostaglandin-H2 D-isomerase | -0.82 | 0.3 | -2.5 | 1.3E-02 | 2.9E-01 | 1.0E+00 |
| MPNI-co17p | P48740 | MASP1 | Mannan-binding lectin serine protease 1 | 0.67 | 0.3 | 2.1 | 3.7E-02 | 3.6E-01 | 1.0E+00 |
| MPNI-co17p | P49747 | COMP | Cartilage oligomeric matrix protein | -0.72 | 0.3 | -2.2 | 3.2E-02 | 3.6E-01 | 1.0E+00 |
| MPNI-co17p | P54289 | CACNA2D1 | Voltage-dependent calcium channel subunit alpha-2/delta-1 | -0.79 | 0.3 | -2.4 | 1.5E-02 | 2.9E-01 | 1.0E+00 |
| MPNI-co17p | P54802 | NAGLU | Alpha-N-acetylglucosaminidase | 0.61 | 0.3 | 2.0 | 4.9E-02 | 4.1E-01 | 1.0E+00 |
| MPNI-co17p | P55290 | CDH13 | Cadherin-13 | -0.68 | 0.3 | -2.1 | 3.6E-02 | 3.6E-01 | 1.0E+00 |
| MPNI-co17p | P68363 | TUBA1B | Tubulin alpha-1B chain | -0.79 | 0.3 | -2.5 | 1.2E-02 | 2.9E-01 | 1.0E+00 |
| MPNI-co17p | P78509 | RELN | Reelin | -0.81 | 0.3 | -2.6 | 9.6E-03 | 2.9E-01 | 1.0E+00 |
| MPNI-co17p | P98160 | HSPG2 | Basement membrane-specific heparan sulfate proteoglycan core protein | -0.64 | 0.3 | -2.0 | 4.6E-02 | 4.0E-01 | 1.0E+00 |
| MPNI-co17p | Q12860 | CNTN1 | Contactin-1 | -0.91 | 0.3 | -2.9 | 4.4E-03 | 2.4E-01 | 1.0E+00 |
| MPNI-co17p | Q14515 | SPARCL1 | SPARC-like protein 1 | -0.92 | 0.3 | -2.9 | 3.4E-03 | 2.1E-01 | 1.0E+00 |
| MPNI-co17p | Q15828 | CST6 | Cystatin-M | -0.72 | 0.3 | -2.2 | 3.0E-02 | 3.6E-01 | 1.0E+00 |
| MPNI-co17p | Q6UX71 | PLXDC2 | Plexin domain-containing protein 2 | -0.71 | 0.3 | -2.1 | 3.3E-02 | 3.6E-01 | 1.0E+00 |
| MPNI-co17p | Q6YHK3 | CD109 | CD109 antigen | -0.89 | 0.3 | -2.8 | 4.8E-03 | 2.4E-01 | 1.0E+00 |
| MPNI-co17p | Q7Z7M0 | MEGF8 | Multiple epidermal growth factor-like domains protein 8 | -0.79 | 0.3 | -2.4 | 1.8E-02 | 3.3E-01 | 1.0E+00 |
| MPNI-co17p | Q86TH1 | ADAMTSL2 | ADAMTS-like protein 2 | -0.71 | 0.3 | -2.2 | 2.6E-02 | 3.6E-01 | 1.0E+00 |
| MPNI-co17p | Q99784 | OLFM1 | Noelin | -0.71 | 0.3 | -2.3 | 2.3E-02 | 3.6E-01 | 1.0E+00 |
| MPNI-co17p | Q99969 | RARRES2 | Retinoic acid receptor responder protein 2 | 0.98 | 0.3 | 3.2 | 1.5E-03 | 1.7E-01 | 6.8E-01 |
| MPNI-co17p | Q9H4B7 | TUBB1 | Tubulin beta-1 chain | -0.66 | 0.3 | -2.1 | 3.7E-02 | 3.6E-01 | 1.0E+00 |
| MPNI-co17p | Q9NPH3 | IL1RAP | Interleukin-1 receptor accessory protein | -0.74 | 0.3 | -2.2 | 3.0E-02 | 3.6E-01 | 1.0E+00 |
| MPNI-co17p | Q9UGM5 | FETUB | Fetuin-B | 0.74 | 0.4 | 2.1 | 3.5E-02 | 3.6E-01 | 1.0E+00 |
| MPNI-co17p | Q9Y5Y7 | LYVE1 | Lymphatic vessel endothelial hyaluronic acid receptor 1 | -0.85 | 0.3 | -2.7 | 7.1E-03 | 2.6E-01 | 1.0E+00 |
| MPNI-co17p | Q9Y6R7 | FCGBP | IgGFc-binding protein | -0.99 | 0.3 | -3.0 | 3.2E-03 | 2.1E-01 | 1.0E+00 |
| MPNI-p12d | P00747 | PLG | Plasminogen | -0.04 | 0.0 | -2.2 | 2.9E-02 | 9.3E-01 | 1.0E+00 |
| MPNI-p12d | P00748 | F12 | Coagulation factor XII | -0.04 | 0.0 | -2.3 | 2.4E-02 | 9.3E-01 | 1.0E+00 |
| MPNI-p12d | P07360 | C8G | Complement component C8 gamma chain | 0.04 | 0.0 | 2.3 | 2.5E-02 | 9.3E-01 | 1.0E+00 |
| MPNI-p12d | P09172 | DBH | Dopamine beta-hydroxylase | -0.05 | 0.0 | -2.7 | 7.2E-03 | 8.5E-01 | 1.0E+00 |
| MPNI-p12d | P10646 | TFPI | Tissue factor pathway inhibitor | 0.04 | 0.0 | 2.3 | 2.4E-02 | 9.3E-01 | 1.0E+00 |
| MPNI-p12d | P11142 | HSPA8 | Heat shock cognate 71 kDa protein | 0.03 | 0.0 | 2.0 | 4.7E-02 | 9.4E-01 | 1.0E+00 |
| MPNI-p12d | P20023 | CR2 | Complement receptor type 2 | 0.04 | 0.0 | 2.2 | 3.1E-02 | 9.3E-01 | 1.0E+00 |
| MPNI-p12d | P24043 | LAMA2 | Laminin subunit alpha-2 | -0.03 | 0.0 | -2.0 | 4.4E-02 | 9.4E-01 | 1.0E+00 |

| | | | | | | | | | |
|---|---|---|---|---|---|---|---|---|---|
| MPNI-p12d | P35908 | KRT2 | Keratin, type II cytoskeletal 2 epidermal | -0.04 | 0.0 | -2.7 | 7.0E-03 | 8.5E-01 | 1.0E+00 |
| MPNI-p12d | P39060 | COL18A1 | Collagen alpha-1(XVIII) chain | 0.04 | 0.0 | 2.2 | 3.2E-02 | 9.3E-01 | 1.0E+00 |
| MPNI-p12d | P41222 | PTGDS | Prostaglandin-H2 D-isomerase | 0.04 | 0.0 | 2.1 | 3.4E-02 | 9.4E-01 | 1.0E+00 |
| MPNI-p12d | P80108 | GPLD1 | Phosphatidylinositol-glycan-specific phospholipase D | -0.03 | 0.0 | -2.0 | 4.3E-02 | 9.4E-01 | 1.0E+00 |
| MPNI-p12d | P80723 | BASP1 | Brain acid soluble protein 1 | 0.04 | 0.0 | 2.5 | 1.4E-02 | 9.3E-01 | 1.0E+00 |
| MPNI-p12d | Q13790 | APOF | Apolipoprotein F | -0.03 | 0.0 | -2.0 | 4.8E-02 | 9.4E-01 | 1.0E+00 |
| MPNI-p12d | Q86SQ4 | ADGRG6 | Adhesion G-protein coupled receptor G6 | 0.04 | 0.0 | 2.2 | 2.9E-02 | 9.3E-01 | 1.0E+00 |
| MPNI-p12d | Q96IY4 | CPB2 | Carboxypeptidase B2 | -0.04 | 0.0 | -2.4 | 1.9E-02 | 9.3E-01 | 1.0E+00 |
| MPNI-p12d | Q96PD5 | PGLYRP2 | N-acetylmuramoyl-L-alanine amidase | 0.05 | 0.0 | 2.6 | 1.1E-02 | 9.3E-01 | 1.0E+00 |
| MPNI-p12d | Q9Y251 | HPSE | Heparanase | -0.04 | 0.0 | -2.7 | 7.8E-03 | 8.5E-01 | 1.0E+00 |
| MPNI-p12d | Q9Y5Y7 | LYVE1 | Lymphatic vessel endothelial hyaluronic acid receptor 1 | -0.05 | 0.0 | -2.8 | 5.1E-03 | 8.5E-01 | 1.0E+00 |
| MPNI-p12d | Q9Y646 | CPQ | Carboxypeptidase Q | 0.03 | 0.0 | 2.2 | 2.9E-02 | 9.3E-01 | 1.0E+00 |
| MPNI-s14d | O00391 | QSOX1 | Sulfhydryl oxidase 1 | -0.05 | 0.0 | -2.8 | 4.6E-03 | 1.4E-01 | 1.0E+00 |
| MPNI-s14d | O00592 | PODXL | Podocalyxin | -0.05 | 0.0 | -2.9 | 4.4E-03 | 1.4E-01 | 1.0E+00 |
| MPNI-s14d | P02452 | COL1A1 | Collagen alpha-1(I) chain | -0.03 | 0.0 | -2.0 | 4.2E-02 | 3.5E-01 | 1.0E+00 |
| MPNI-s14d | P02743 | APCS | Serum amyloid P-component | 0.05 | 0.0 | 2.9 | 3.6E-03 | 1.4E-01 | 1.0E+00 |
| MPNI-s14d | P02747 | C1QC | Complement C1q subcomponent subunit C | -0.03 | 0.0 | -2.2 | 2.5E-02 | 3.1E-01 | 1.0E+00 |
| MPNI-s14d | P04004 | VTN | Vitronectin | 0.03 | 0.0 | 2.1 | 3.7E-02 | 3.5E-01 | 1.0E+00 |
| MPNI-s14d | P04066 | FUCA1 | Tissue alpha-L-fucosidase | -0.03 | 0.0 | -2.1 | 4.0E-02 | 3.5E-01 | 1.0E+00 |
| MPNI-s14d | P04196 | HRG | Histidine-rich glycoprotein | -0.04 | 0.0 | -2.6 | 8.5E-03 | 2.0E-01 | 1.0E+00 |
| MPNI-s14d | P05090 | APOD | Apolipoprotein D | -0.03 | 0.0 | -2.0 | 4.1E-02 | 3.5E-01 | 1.0E+00 |
| MPNI-s14d | P05543 | SERPINA7 | Thyroxine-binding globulin | 0.04 | 0.0 | 2.2 | 3.2E-02 | 3.4E-01 | 1.0E+00 |
| MPNI-s14d | P06396 | GSN | Gelsolin | -0.04 | 0.0 | -2.5 | 1.3E-02 | 2.4E-01 | 1.0E+00 |
| MPNI-s14d | P08253 | MMP2 | 72 kDa type IV collagenase | -0.04 | 0.0 | -2.8 | 5.4E-03 | 1.5E-01 | 1.0E+00 |
| MPNI-s14d | P0C0L4 | C4A | Complement C4-A | 0.05 | 0.0 | 3.1 | 2.0E-03 | 1.2E-01 | 8.7E-01 |
| MPNI-s14d | P0C0L5 | C4B | Complement C4-B | 0.04 | 0.0 | 2.8 | 5.9E-03 | 1.5E-01 | 1.0E+00 |
| MPNI-s14d | P11362 | FGFR1 | Fibroblast growth factor receptor 1 | -0.03 | 0.0 | -2.1 | 4.1E-02 | 3.5E-01 | 1.0E+00 |
| MPNI-s14d | P12259 | F5 | Coagulation factor V | -0.03 | 0.0 | -2.1 | 3.6E-02 | 3.5E-01 | 1.0E+00 |
| MPNI-s14d | P12955 | PEPD | Xaa-Pro dipeptidase | -0.03 | 0.0 | -2.3 | 2.1E-02 | 3.1E-01 | 1.0E+00 |
| MPNI-s14d | P13598 | ICAM2 | Intercellular adhesion molecule 2 | -0.03 | 0.0 | -2.2 | 3.0E-02 | 3.4E-01 | 1.0E+00 |
| MPNI-s14d | P13796 | LCP1 | Plastin-2 | -0.04 | 0.0 | -2.5 | 1.4E-02 | 2.4E-01 | 1.0E+00 |
| MPNI-s14d | P14618 | PKM | Pyruvate kinase PKM | -0.04 | 0.0 | -2.8 | 4.9E-03 | 1.4E-01 | 1.0E+00 |
| MPNI-s14d | P16035 | TIMP2 | Metalloproteinase inhibitor 2 | -0.04 | 0.0 | -2.6 | 1.0E-02 | 2.3E-01 | 1.0E+00 |
| MPNI-s14d | P22105 | TNXB | Tenascin-X | -0.04 | 0.0 | -2.8 | 4.5E-03 | 1.4E-01 | 1.0E+00 |

| | | | | | | | | | |
|---|---|---|---|---|---|---|---|---|---|
| MPNI-s14d | P24593 | IGFBP5 | Insulin-like growth factor-binding protein 5 | -0.05 | 0.0 | -3.1 | 2.3E-03 | 1.2E-01 | 9.9E-01 |
| MPNI-s14d | P24821 | TNC | Tenascin | -0.05 | 0.0 | -3.2 | 1.5E-03 | 1.2E-01 | 6.6E-01 |
| MPNI-s14d | P33151 | CDH5 | Cadherin-5 | -0.04 | 0.0 | -2.4 | 1.8E-02 | 2.7E-01 | 1.0E+00 |
| MPNI-s14d | P35443 | THBS4 | Thrombospondin-4 | -0.04 | 0.0 | -2.3 | 2.2E-02 | 3.1E-01 | 1.0E+00 |
| MPNI-s14d | P39060 | COL18A1 | Collagen alpha-1(XVIII) chain | -0.03 | 0.0 | -2.2 | 2.8E-02 | 3.4E-01 | 1.0E+00 |
| MPNI-s14d | P40189 | IL6ST | Interleukin-6 receptor subunit beta | -0.04 | 0.0 | -3.0 | 3.0E-03 | 1.4E-01 | 1.0E+00 |
| MPNI-s14d | P43121 | MCAM | Cell surface glycoprotein MUC18 | -0.04 | 0.0 | -2.5 | 1.4E-02 | 2.4E-01 | 1.0E+00 |
| MPNI-s14d | P49747 | COMP | Cartilage oligomeric matrix protein | -0.04 | 0.0 | -2.2 | 2.9E-02 | 3.4E-01 | 1.0E+00 |
| MPNI-s14d | P54289 | CACNA2D1 | Voltage-dependent calcium channel subunit alpha-2/delta-1 | -0.05 | 0.0 | -3.2 | 1.5E-03 | 1.2E-01 | 6.7E-01 |
| MPNI-s14d | P55290 | CDH13 | Cadherin-13 | -0.05 | 0.0 | -3.4 | 8.5E-04 | 1.2E-01 | 3.7E-01 |
| MPNI-s14d | P80723 | BASP1 | Brain acid soluble protein 1 | -0.03 | 0.0 | -2.0 | 5.0E-02 | 4.0E-01 | 1.0E+00 |
| MPNI-s14d | P98160 | HSPG2 | Basement membrane-specific heparan sulfate proteoglycan core protein | -0.03 | 0.0 | -2.2 | 2.8E-02 | 3.4E-01 | 1.0E+00 |
| MPNI-s14d | Q04756 | HGFAC | Hepatocyte growth factor activator | -0.03 | 0.0 | -2.0 | 4.2E-02 | 3.5E-01 | 1.0E+00 |
| MPNI-s14d | Q07954 | LRP1 | Prolow-density lipoprotein receptor-related protein 1 | -0.03 | 0.0 | -2.3 | 2.4E-02 | 3.1E-01 | 1.0E+00 |
| MPNI-s14d | Q10588 | BST1 | ADP-ribosyl cyclase/cyclic ADP-ribose hydrolase 2 | -0.04 | 0.0 | -2.7 | 6.1E-03 | 1.5E-01 | 1.0E+00 |
| MPNI-s14d | Q12860 | CNTN1 | Contactin-1 | -0.05 | 0.0 | -3.3 | 8.9E-04 | 1.2E-01 | 3.9E-01 |
| MPNI-s14d | Q13332 | PTPRS | Receptor-type tyrosine-protein phosphatase S | -0.04 | 0.0 | -2.9 | 3.8E-03 | 1.4E-01 | 1.0E+00 |
| MPNI-s14d | Q13449 | LSAMP | Limbic system-associated membrane protein | -0.03 | 0.0 | -2.2 | 3.2E-02 | 3.4E-01 | 1.0E+00 |
| MPNI-s14d | Q14515 | SPARCL1 | SPARC-like protein 1 | -0.04 | 0.0 | -2.4 | 1.7E-02 | 2.7E-01 | 1.0E+00 |
| MPNI-s14d | Q14956 | GPNMB | Transmembrane glycoprotein NMB | -0.05 | 0.0 | -3.2 | 1.7E-03 | 1.2E-01 | 7.4E-01 |
| MPNI-s14d | Q6EMK4 | VASN | Vasorin | -0.03 | 0.0 | -2.1 | 4.0E-02 | 3.5E-01 | 1.0E+00 |
| MPNI-s14d | Q6UX71 | PLXDC2 | Plexin domain-containing protein 2 | -0.04 | 0.0 | -2.5 | 1.1E-02 | 2.4E-01 | 1.0E+00 |
| MPNI-s14d | Q6UY14 | ADAMTSL4 | ADAMTS-like protein 4 | -0.04 | 0.0 | -2.5 | 1.3E-02 | 2.4E-01 | 1.0E+00 |
| MPNI-s14d | Q6YHK3 | CD109 | CD109 antigen | -0.04 | 0.0 | -2.5 | 1.3E-02 | 2.4E-01 | 1.0E+00 |
| MPNI-s14d | Q7Z7M0 | MEGF8 | Multiple epidermal growth factor-like domains protein 8 | -0.03 | 0.0 | -2.1 | 3.4E-02 | 3.5E-01 | 1.0E+00 |
| MPNI-s14d | Q86U17 | SERPINA11 | Serpin A11 | -0.03 | 0.0 | -2.1 | 3.8E-02 | 3.5E-01 | 1.0E+00 |
| MPNI-s14d | Q8IUL8 | CILP2 | Cartilage intermediate layer protein 2 | -0.03 | 0.0 | -2.3 | 2.4E-02 | 3.1E-01 | 1.0E+00 |
| MPNI-s14d | Q8WZ75 | ROBO4 | Roundabout homolog 4 | -0.04 | 0.0 | -2.4 | 1.6E-02 | 2.5E-01 | 1.0E+00 |
| MPNI-s14d | Q96NZ9 | PRAP1 | Proline-rich acidic protein 1 | -0.03 | 0.0 | -2.1 | 4.1E-02 | 3.5E-01 | 1.0E+00 |
| MPNI-s14d | Q99983 | OMD | Osteomodulin | -0.05 | 0.0 | -3.2 | 1.7E-03 | 1.2E-01 | 7.4E-01 |
| MPNI-s14d | Q9NY15 | STAB1 | Stabilin-1 | -0.03 | 0.0 | -2.3 | 2.3E-02 | 3.1E-01 | 1.0E+00 |

| | | | | | | | | | |
|---|---|---|---|---|---|---|---|---|---|
| MPNI-s14d | Q9UK55 | SERPINA10 | Protein Z-dependent protease inhibitor | 0.03 | 0.0 | 2.1 | 3.7E-02 | 3.5E-01 | 1.0E+00 |
| MPNI-s17d | O43493 | TGOLN2 | Trans-Golgi network integral membrane protein 2 | 0.06 | 0.0 | 2.1 | 3.5E-02 | 6.7E-01 | 1.0E+00 |
| MPNI-s17d | O75594 | PGLYRP1 | Peptidoglycan recognition protein 1 | -0.06 | 0.0 | -2.2 | 2.8E-02 | 6.7E-01 | 1.0E+00 |
| MPNI-s17d | P00747 | PLG | Plasminogen | -0.06 | 0.0 | -2.2 | 3.1E-02 | 6.7E-01 | 1.0E+00 |
| MPNI-s17d | P02743 | APCS | Serum amyloid P-component | 0.06 | 0.0 | 2.2 | 2.9E-02 | 6.7E-01 | 1.0E+00 |
| MPNI-s17d | P04003 | C4BPA | C4b-binding protein alpha chain | 0.06 | 0.0 | 2.2 | 2.9E-02 | 6.7E-01 | 1.0E+00 |
| MPNI-s17d | P05062 | ALDOB | Fructose-bisphosphate aldolase B | 0.08 | 0.0 | 3.0 | 3.1E-03 | 6.7E-01 | 1.0E+00 |
| MPNI-s17d | P06727 | APOA4 | Apolipoprotein A-IV | -0.06 | 0.0 | -2.1 | 3.7E-02 | 6.7E-01 | 1.0E+00 |
| MPNI-s17d | P07360 | C8G | Complement component C8 gamma chain | 0.06 | 0.0 | 2.1 | 3.5E-02 | 6.7E-01 | 1.0E+00 |
| MPNI-s17d | P08195 | SLC3A2 | 4F2 cell-surface antigen heavy chain | 0.06 | 0.0 | 2.1 | 3.6E-02 | 6.7E-01 | 1.0E+00 |
| MPNI-s17d | P08238 | HSP90AB1 | Heat shock protein HSP 90-beta | 0.08 | 0.0 | 2.9 | 3.7E-03 | 6.7E-01 | 1.0E+00 |
| MPNI-s17d | P08294 | SOD3 | Extracellular superoxide dismutase [Cu-Zn] | 0.08 | 0.0 | 2.8 | 5.4E-03 | 6.7E-01 | 1.0E+00 |
| MPNI-s17d | P10153 | RNASE2 | Non-secretory ribonuclease | -0.05 | 0.0 | -2.0 | 4.6E-02 | 7.3E-01 | 1.0E+00 |
| MPNI-s17d | P10721 | KIT | Mast/stem cell growth factor receptor Kit | -0.06 | 0.0 | -2.3 | 2.3E-02 | 6.7E-01 | 1.0E+00 |
| MPNI-s17d | P11362 | FGFR1 | Fibroblast growth factor receptor 1 | -0.07 | 0.0 | -2.5 | 1.2E-02 | 6.7E-01 | 1.0E+00 |
| MPNI-s17d | P12955 | PEPD | Xaa-Pro dipeptidase | -0.07 | 0.0 | -2.7 | 8.1E-03 | 6.7E-01 | 1.0E+00 |
| MPNI-s17d | P13671 | C6 | Complement component C6 | 0.06 | 0.0 | 2.3 | 2.2E-02 | 6.7E-01 | 1.0E+00 |
| MPNI-s17d | P14625 | HSP90B1 | Endoplasmin | 0.06 | 0.0 | 2.1 | 3.6E-02 | 6.7E-01 | 1.0E+00 |
| MPNI-s17d | P15151 | PVR | Poliovirus receptor | -0.05 | 0.0 | -2.0 | 4.3E-02 | 7.3E-01 | 1.0E+00 |
| MPNI-s17d | P17936 | IGFBP3 | Insulin-like growth factor-binding protein 3 | 0.05 | 0.0 | 2.0 | 4.7E-02 | 7.3E-01 | 1.0E+00 |
| MPNI-s17d | P19022 | CDH2 | Cadherin-2 | 0.06 | 0.0 | 2.4 | 1.8E-02 | 6.7E-01 | 1.0E+00 |
| MPNI-s17d | P29279 | CCN2 | CCN family member 2 | -0.06 | 0.0 | -2.2 | 2.5E-02 | 6.7E-01 | 1.0E+00 |
| MPNI-s17d | P32942 | ICAM3 | Intercellular adhesion molecule 3 | -0.07 | 0.0 | -2.6 | 1.0E-02 | 6.7E-01 | 1.0E+00 |
| MPNI-s17d | P39060 | COL18A1 | Collagen alpha-1(XVIII) chain | 0.06 | 0.0 | 2.3 | 2.3E-02 | 6.7E-01 | 1.0E+00 |
| MPNI-s17d | P53634 | CTSC | Dipeptidyl peptidase 1 | -0.05 | 0.0 | -2.0 | 5.0E-02 | 7.5E-01 | 1.0E+00 |
| MPNI-s17d | Q14956 | GPNMB | Transmembrane glycoprotein NMB | -0.06 | 0.0 | -2.4 | 1.7E-02 | 6.7E-01 | 1.0E+00 |
| MPNI-s17d | Q16853 | AOC3 | Membrane primary amine oxidase | 0.06 | 0.0 | 2.1 | 3.3E-02 | 6.7E-01 | 1.0E+00 |
| MPNI-s17d | Q86SQ4 | ADGRG6 | Adhesion G-protein coupled receptor G6 | 0.06 | 0.0 | 2.1 | 3.7E-02 | 6.7E-01 | 1.0E+00 |
| MPNI-s17d | Q9H299 | SH3BGRL3 | SH3 domain-binding glutamic acid-rich-like protein 3 | -0.05 | 0.0 | -2.1 | 3.7E-02 | 6.7E-01 | 1.0E+00 |
| MPNI-s17d | Q9NPR2 | SEMA4B | Semaphorin-4B | 0.06 | 0.0 | 2.1 | 3.9E-02 | 6.8E-01 | 1.0E+00 |
| MPNI-t12d | P00338 | LDHA | L-lactate dehydrogenase A chain | -0.04 | 0.0 | -2.5 | 1.1E-02 | 7.3E-01 | 1.0E+00 |
| MPNI-t12d | P00742 | F10 | Coagulation factor X | 0.04 | 0.0 | 2.2 | 3.1E-02 | 7.7E-01 | 1.0E+00 |

| | | | | | | | | | |
|---|---|---|---|---|---|---|---|---|---|
| MPNI-t12d | P02042 | HBD | Hemoglobin subunit delta | -0.04 | 0.0 | -2.3 | 2.0E-02 | 7.3E-01 | 1.0E+00 |
| MPNI-t12d | P04439 | HLA-A | HLA class I histocompatibility antigen, A alpha chain | -0.07 | 0.0 | -3.2 | 1.5E-03 | 3.3E-01 | 6.5E-01 |
| MPNI-t12d | P04746 | AMY2A | Pancreatic alpha-amylase | -0.05 | 0.0 | -2.7 | 7.2E-03 | 7.3E-01 | 1.0E+00 |
| MPNI-t12d | P06732 | CKM | Creatine kinase M-type | -0.04 | 0.0 | -2.1 | 3.4E-02 | 7.8E-01 | 1.0E+00 |
| MPNI-t12d | P07195 | LDHB | L-lactate dehydrogenase B chain | -0.04 | 0.0 | -2.4 | 1.5E-02 | 7.3E-01 | 1.0E+00 |
| MPNI-t12d | P07307 | ASGR2 | Asialoglycoprotein receptor 2 | -0.04 | 0.0 | -2.3 | 2.4E-02 | 7.7E-01 | 1.0E+00 |
| MPNI-t12d | P11362 | FGFR1 | Fibroblast growth factor receptor 1 | -0.04 | 0.0 | -2.6 | 1.0E-02 | 7.3E-01 | 1.0E+00 |
| MPNI-t12d | P12109 | COL6A1 | Collagen alpha-1(VI) chain | -0.04 | 0.0 | -2.4 | 1.8E-02 | 7.3E-01 | 1.0E+00 |
| MPNI-t12d | P22897 | MRC1 | Macrophage mannose receptor 1 | -0.04 | 0.0 | -2.4 | 1.6E-02 | 7.3E-01 | 1.0E+00 |
| MPNI-t12d | P23470 | PTPRG | Receptor-type tyrosine-protein phosphatase gamma | 0.05 | 0.0 | 2.5 | 1.3E-02 | 7.3E-01 | 1.0E+00 |
| MPNI-t12d | P35858 | IGFALS | Insulin-like growth factor-binding protein complex acid labile subunit | 0.04 | 0.0 | 2.0 | 4.8E-02 | 9.5E-01 | 1.0E+00 |
| MPNI-t12d | P43652 | AFM | Afamin | 0.04 | 0.0 | 2.2 | 3.1E-02 | 7.7E-01 | 1.0E+00 |
| MPNI-t12d | P61769 | B2M | Beta-2-microglobulin | -0.04 | 0.0 | -2.2 | 3.2E-02 | 7.7E-01 | 1.0E+00 |
| MPNI-t12d | Q06033 | ITIH3 | Inter-alpha-trypsin inhibitor heavy chain H3 | -0.05 | 0.0 | -2.4 | 1.6E-02 | 7.3E-01 | 1.0E+00 |
| MPNI-t12d | Q12907 | LMAN2 | Vesicular integral-membrane protein VIP36 | 0.05 | 0.0 | 3.2 | 1.4E-03 | 3.3E-01 | 6.3E-01 |
| MPNI-t12d | Q9NZK5 | ADA2 | Adenosine deaminase 2 | -0.04 | 0.0 | -2.2 | 3.0E-02 | 7.7E-01 | 1.0E+00 |
| MPNI-t12d | Q9UBX1 | CTSF | Cathepsin F | -0.04 | 0.0 | -2.1 | 3.7E-02 | 8.2E-01 | 1.0E+00 |
| MPNI-t12d | Q9UHG3 | PCYOX1 | Prenylcysteine oxidase 1 | -0.04 | 0.0 | -2.2 | 3.0E-02 | 7.7E-01 | 1.0E+00 |
| MPNI-t12d | Q9UK55 | SERPINA10 | Protein Z-dependent protease inhibitor | -0.04 | 0.0 | -2.0 | 4.1E-02 | 8.6E-01 | 1.0E+00 |
| MPNI-t12d | Q9ULI3 | HEG1 | Protein HEG homolog 1 | 0.04 | 0.0 | 2.3 | 2.0E-02 | 7.3E-01 | 1.0E+00 |
| MPNI-t14d | O95497 | VNN1 | Pantetheinase | 0.04 | 0.0 | 2.2 | 3.1E-02 | 9.8E-01 | 1.0E+00 |
| MPNI-t14d | P06681 | C2 | Complement C2 | 0.05 | 0.0 | 2.7 | 7.8E-03 | 9.8E-01 | 1.0E+00 |
| MPNI-t14d | P07602 | PSAP | Prosaposin | 0.05 | 0.0 | 2.5 | 1.2E-02 | 9.8E-01 | 1.0E+00 |
| MPNI-t14d | P11597 | CETP | Cholesteryl ester transfer protein | -0.04 | 0.0 | -2.3 | 2.1E-02 | 9.8E-01 | 1.0E+00 |
| MPNI-t14d | P19021 | PAM | Peptidyl-glycine alpha-amidating monooxygenase | -0.04 | 0.0 | -2.1 | 3.7E-02 | 9.8E-01 | 1.0E+00 |
| MPNI-t14d | P24821 | TNC | Tenascin | 0.04 | 0.0 | 2.2 | 2.8E-02 | 9.8E-01 | 1.0E+00 |
| MPNI-t14d | P33908 | MAN1A1 | Mannosyl-oligosaccharide 1,2-alpha-mannosidase IA | 0.04 | 0.0 | 2.1 | 4.0E-02 | 9.8E-01 | 1.0E+00 |
| MPNI-t14d | Q08380 | LGALS3BP | Galectin-3-binding protein | -0.05 | 0.0 | -2.2 | 2.9E-02 | 9.8E-01 | 1.0E+00 |
| MPNI-t14d | Q9HDC9 | APMAP | Adipocyte plasma membrane-associated protein | -0.04 | 0.0 | -2.0 | 4.2E-02 | 9.8E-01 | 1.0E+00 |
| MPNI-t14d | Q9NTU7 | CBLN4 | Cerebellin-4 | 0.04 | 0.0 | 2.0 | 5.0E-02 | 9.8E-01 | 1.0E+00 |
| MPNI-t14d | Q9UNN8 | PROCR | Endothelial protein C receptor | -0.05 | 0.0 | -2.4 | 1.5E-02 | 9.8E-01 | 1.0E+00 |

| | | | | | | | | | |
|---|---|---|---|---|---|---|---|---|---|
| MPNI-co14d | O75594 | PGLYRP1 | Peptidoglycan recognition protein 1 | -0.03 | 0.0 | -2.0 | 5.0E-02 | 8.9E-01 | 1.0E+00 |
| MPNI-co14d | P00533 | EGFR | Epidermal growth factor receptor | -0.03 | 0.0 | -2.0 | 4.8E-02 | 8.9E-01 | 1.0E+00 |
| MPNI-co14d | P00734 | F2 | Prothrombin | 0.03 | 0.0 | 2.0 | 4.7E-02 | 8.9E-01 | 1.0E+00 |
| MPNI-co14d | P02649 | APOE | Apolipoprotein E | 0.03 | 0.0 | 2.1 | 3.2E-02 | 8.9E-01 | 1.0E+00 |
| MPNI-co14d | P02654 | APOC1 | Apolipoprotein C-I | 0.03 | 0.0 | 2.0 | 4.7E-02 | 8.9E-01 | 1.0E+00 |
| MPNI-co14d | P02743 | APCS | Serum amyloid P-component | 0.05 | 0.0 | 2.7 | 7.0E-03 | 7.6E-01 | 1.0E+00 |
| MPNI-co14d | P05362 | ICAM1 | Intercellular adhesion molecule 1 | 0.04 | 0.0 | 2.5 | 1.1E-02 | 8.9E-01 | 1.0E+00 |
| MPNI-co14d | P08493 | MGP | Matrix Gla protein | -0.04 | 0.0 | -2.2 | 2.8E-02 | 8.9E-01 | 1.0E+00 |
| MPNI-co14d | P09960 | LTA4H | Leukotriene A-4 hydrolase | 0.03 | 0.0 | 2.1 | 3.5E-02 | 8.9E-01 | 1.0E+00 |
| MPNI-co14d | P0C0L5 | C4B | Complement C4-B | 0.04 | 0.0 | 2.3 | 2.2E-02 | 8.9E-01 | 1.0E+00 |
| MPNI-co14d | P13598 | ICAM2 | Intercellular adhesion molecule 2 | -0.03 | 0.0 | -2.1 | 3.7E-02 | 8.9E-01 | 1.0E+00 |
| MPNI-co14d | P16070 | CD44 | CD44 antigen | 0.03 | 0.0 | 2.1 | 3.9E-02 | 8.9E-01 | 1.0E+00 |
| MPNI-co14d | P20851 | C4BPB | C4b-binding protein beta chain | 0.05 | 0.0 | 2.7 | 6.8E-03 | 7.6E-01 | 1.0E+00 |
| MPNI-co14d | P23470 | PTPRG | Receptor-type tyrosine-protein phosphatase gamma | 0.05 | 0.0 | 3.0 | 3.1E-03 | 7.6E-01 | 1.0E+00 |
| MPNI-co14d | P35527 | KRT9 | Keratin, type I cytoskeletal 9 | 0.04 | 0.0 | 2.4 | 1.9E-02 | 8.9E-01 | 1.0E+00 |
| MPNI-co14d | P35542 | SAA4 | Serum amyloid A-4 protein | 0.04 | 0.0 | 2.2 | 2.9E-02 | 8.9E-01 | 1.0E+00 |
| MPNI-co14d | P35555 | FBN1 | Fibrillin-1 | 0.03 | 0.0 | 2.0 | 4.7E-02 | 8.9E-01 | 1.0E+00 |
| MPNI-co14d | Q14314 | FGL2 | Fibroleukin | -0.04 | 0.0 | -2.1 | 3.5E-02 | 8.9E-01 | 1.0E+00 |
| MPNI-co14d | Q5D862 | FLG2 | Filaggrin-2 | 0.03 | 0.0 | 2.0 | 4.6E-02 | 8.9E-01 | 1.0E+00 |
| MPNI-co14d | Q6UXB8 | PI16 | Peptidase inhibitor 16 | 0.04 | 0.0 | 2.4 | 1.5E-02 | 8.9E-01 | 1.0E+00 |
| MPNI-co14d | Q86SQ4 | ADGRG6 | Adhesion G-protein coupled receptor G6 | 0.04 | 0.0 | 2.3 | 2.2E-02 | 8.9E-01 | 1.0E+00 |
| MPNI-co14d | Q92954 | PRG4 | Proteoglycan 4 | 0.04 | 0.0 | 2.4 | 1.9E-02 | 8.9E-01 | 1.0E+00 |
| MPNI-co14d | Q96NZ9 | PRAP1 | Proline-rich acidic protein 1 | -0.05 | 0.0 | -2.9 | 3.6E-03 | 7.6E-01 | 1.0E+00 |
| MPNI-co17d | A1L4H1 | SSC5D | Soluble scavenger receptor cysteine-rich domain-containing protein SSC5D | 0.05 | 0.0 | 2.0 | 4.8E-02 | 9.4E-01 | 1.0E+00 |
| MPNI-co17d | O15204 | ADAMDEC1 | ADAM DEC1 | -0.07 | 0.0 | -2.9 | 3.3E-03 | 7.5E-01 | 1.0E+00 |
| MPNI-co17d | O75594 | PGLYRP1 | Peptidoglycan recognition protein 1 | -0.07 | 0.0 | -2.7 | 6.2E-03 | 7.5E-01 | 1.0E+00 |
| MPNI-co17d | P00747 | PLG | Plasminogen | -0.05 | 0.0 | -2.0 | 4.9E-02 | 9.4E-01 | 1.0E+00 |
| MPNI-co17d | P01011 | SERPINA3 | Alpha-1-antichymotrypsin | 0.05 | 0.0 | 2.1 | 3.9E-02 | 9.1E-01 | 1.0E+00 |
| MPNI-co17d | P01031 | C5 | Complement C5 | 0.05 | 0.0 | 2.1 | 3.9E-02 | 9.1E-01 | 1.0E+00 |
| MPNI-co17d | P06727 | APOA4 | Apolipoprotein A-IV | -0.05 | 0.0 | -2.0 | 4.6E-02 | 9.4E-01 | 1.0E+00 |
| MPNI-co17d | P07911 | UMOD | Uromodulin | -0.05 | 0.0 | -2.1 | 3.4E-02 | 9.1E-01 | 1.0E+00 |
| MPNI-co17d | P08514 | ITGA2B | Integrin alpha-IIb | -0.06 | 0.0 | -2.4 | 1.9E-02 | 7.5E-01 | 1.0E+00 |
| MPNI-co17d | P12830 | CDH1 | Cadherin-1 | -0.06 | 0.0 | -2.4 | 1.5E-02 | 7.5E-01 | 1.0E+00 |
| MPNI-co17d | P13646 | KRT13 | Keratin, type I cytoskeletal 13 | 0.06 | 0.0 | 2.4 | 1.5E-02 | 7.5E-01 | 1.0E+00 |
| MPNI-co17d | P13671 | C6 | Complement component C6 | 0.06 | 0.0 | 2.3 | 2.1E-02 | 7.8E-01 | 1.0E+00 |
| MPNI-co17d | P14543 | NID1 | Nidogen-1 | -0.05 | 0.0 | -2.1 | 3.9E-02 | 9.1E-01 | 1.0E+00 |

| | | | | | | | | | |
|---|---|---|---|---|---|---|---|---|---|
| MPNI-co17d | P22692 | IGFBP4 | Insulin-like growth factor-binding protein 4 | -0.06 | 0.0 | -2.5 | 1.3E-02 | 7.5E-01 | 1.0E+00 |
| MPNI-co17d | P55290 | CDH13 | Cadherin-13 | -0.07 | 0.0 | -2.7 | 7.7E-03 | 7.5E-01 | 1.0E+00 |
| MPNI-co17d | Q10588 | BST1 | ADP-ribosyl cyclase/cyclic ADP-ribose hydrolase 2 | -0.06 | 0.0 | -2.4 | 1.8E-02 | 7.5E-01 | 1.0E+00 |
| MPNI-co17d | Q13093 | PLA2G7 | Platelet-activating factor acetylhydrolase | 0.06 | 0.0 | 2.2 | 2.7E-02 | 9.1E-01 | 1.0E+00 |
| MPNI-co17d | Q86SQ4 | ADGRG6 | Adhesion G-protein coupled receptor G6 | 0.07 | 0.0 | 2.6 | 1.1E-02 | 7.5E-01 | 1.0E+00 |
| MPNI-co17d | Q9H299 | SH3BGRL3 | SH3 domain-binding glutamic acid-rich-like protein 3 | -0.06 | 0.0 | -2.4 | 1.8E-02 | 7.5E-01 | 1.0E+00 |
| MPNI-co17d | Q9HDC9 | APMAP | Adipocyte plasma membrane-associated protein | -0.06 | 0.0 | -2.5 | 1.2E-02 | 7.5E-01 | 1.0E+00 |
| MPNI-co17d | Q9NZK5 | ADA2 | Adenosine deaminase 2 | -0.05 | 0.0 | -2.1 | 3.6E-02 | 9.1E-01 | 1.0E+00 |
| MPNI-co17d | Q9UJJ9 | GNPTG | N-acetylglucosamine-1-phosphotransferase subunit gamma | -0.05 | 0.0 | -2.1 | 3.9E-02 | 9.1E-01 | 1.0E+00 |
